# Inherited variants in *CHD3* demonstrate variable expressivity in Snijders Blok-Campeau syndrome

**DOI:** 10.1101/2021.10.04.21264162

**Authors:** Jet van der Spek, Joery den Hoed, Lot Snijders Blok, Alexander J. M. Dingemans, Dick Schijven, Christoffer Nellaker, Hanka Venselaar, Tahsin Stefan Barakat, E. Martina Bebin, Stefanie Beck-Wödl, Gea Beunders, Natasha J. Brown, Theresa Brunet, Han G. Brunner, Philippe M. Campeau, Goran Čuturilo, Christian Gilissen, Tobias B. Haack, Ralf A. Husain, Benjamin Kamien, Sze Chern Lim, Luca Lovrecic, Janine Magg, Ales Maver, Valancy Miranda, Danielle C. Monteil, Charlotte W. Ockeloen, Lynn S. Pais, Vasilica Plaiasu, Laura Raiti, Christopher Richmond, Angelika Rieß, Eva M. C. Schwaibold, Marleen E. H. Simon, Stephanie Spranger, Tiong Yang Tan, Michelle L. Thompson, Bert B.A. de Vries, Ella J. Wilkins, Marjolein H. Willemsen, Clyde Francks, Lisenka E. L. M. Vissers, Simon E. Fisher, Tjitske Kleefstra

## Abstract

Interpretation of next-generation sequencing data of individuals with an apparent sporadic neurodevelopmental disorder (NDD) often focusses on pathogenic variants in genes associated with NDD, assuming full clinical penetrance with limited variable expressivity. Consequently, inherited variants in genes associated with dominant disorders may be overlooked when the transmitting parent is clinically unaffected. While *de novo* variants explain a substantial proportion of cases with NDDs, a significant number remains undiagnosed possibly explained by coding variants associated with reduced penetrance and variable expressivity. We characterized twenty families with inherited heterozygous missense or protein-truncating variants (PTVs) in *CHD3*, a gene in which *de novo* variants cause Snijders Blok-Campeau syndrome, characterized by intellectual disability, speech delay and recognizable facial features (SNIBCPS). Notably, the majority of the inherited *CHD3* variants were maternally transmitted. Computational facial and human phenotype ontology-based comparisons demonstrated that the phenotypic features of probands with inherited *CHD3* variants overlap with the phenotype previously associated with *de novo* variants in the gene, while carrier parents are mildly or not affected, suggesting variable expressivity. Additionally, similarly reduced expression levels of CHD3 protein in cells of an affected proband and of related healthy carriers with a *CHD3* PTV, suggested that compensation of expression from the wildtype allele is unlikely to be an underlying mechanism. Our results point to a significant role of inherited variation in SNIBCPS, a finding that is critical for correct variant interpretation and genetic counseling and warrants further investigation towards understanding the broader contributions of such variation to the landscape of human disease.

## Main text

The availability of whole exome sequencing (WES) in clinical practice has greatly improved the yield of genetic diagnostics for individuals with neurodevelopmental disorders (NDDs). In particular, sequencing of proband-parent trios, followed by filtering for *de novo* ^1-3^ or bi-allelic variants ^4; 5^, has proven a powerful tool to identify causal variants in individuals with sporadic dominant and recessive NDDs. However, while *de novo* and bi-allelic variants explain a substantial proportion of cases with NDDs ^1; 4; 5^, the majority remains undiagnosed ^6^. Various factors may explain the difficulties to diagnose these individuals, including variation in genes not yet associated to disease, polygenic inheritance or variation in non-coding regions ^7^. Also coding variants associated with reduced penetrance and variable expressivity may underlie unexplained NDD cases ^6; 8^. Common diagnostic strategies to analyze next-generation sequencing data are not optimized to identify the contributions of these factors to disease. While penetrance indicates the proportion of carriers of a particular variant with a phenotype, expressivity describes the variability in severity of the phenotype between carriers of this variant ^9^. Variable expressivity can cause highly variable symptoms, even in severe disorders that are caused by variants with a large effect ^9; 10^.

In the present study we show variable expressivity for variation in *CHD3*. CHD3 is an ATP-dependent chromatin remodeling protein that serves as core member of the NuRD complex ^11^. Heterozygous variants in *CHD3* have recently been shown to cause a neurodevelopmental syndrome with a variable phenotype, ranging from mildly to more severely affected cases (MIM #618205, Snijders Blok-Campeau syndrome: SNIBCPS) ^12; 13^. *CHD3* is extremely intolerant for both loss-of-function (LoF) and missense variation (pLI = 1, o/e = 0.09 (0.05 - 0.15); Z = 6.15, o/e = 0.5 (0.46 – 0.53)), suggesting haploinsufficiency as a possible disease mechanism. However, the large majority of cases diagnosed with SNIBCPS carry confirmed *de novo* missense variants or single amino acid in-frame deletion variants (51/55, 93% of cases) ^12-14^, clustering in the ATPase-Helicase domain of the encoded protein, and affecting its ATPase activity and/or chromatin remodeling functions, which could be consistent with a dominant-negative mechanism^12^.

Here, we identified twenty families with SNIBCPS, each initially identified through a proband diagnosed with a syndromic NDD carrying a rare inherited *CHD3* missense variant (n = 12) or PTV (n = 8) (NM_001005273.2/ENST00000330494.7; Figure 1). Based on clinical observations, all probands had phenotypes overlapping with the SNIBCPS-phenotype associated with *de novo* variants in *CHD3* (Figure 2; Supplemental Notes 1; Table 1, S1 and S2). Computational facial analysis also confirmed the presence of a SNIBCPS facial gestalt in probands (Figure S3; Table S2), and composite images showed similarities in facial features between probands with *de novo* and inherited *CHD3* variants (squared face, deep set eyes, pointed chin; Figure 2B).

**Figure 1.**
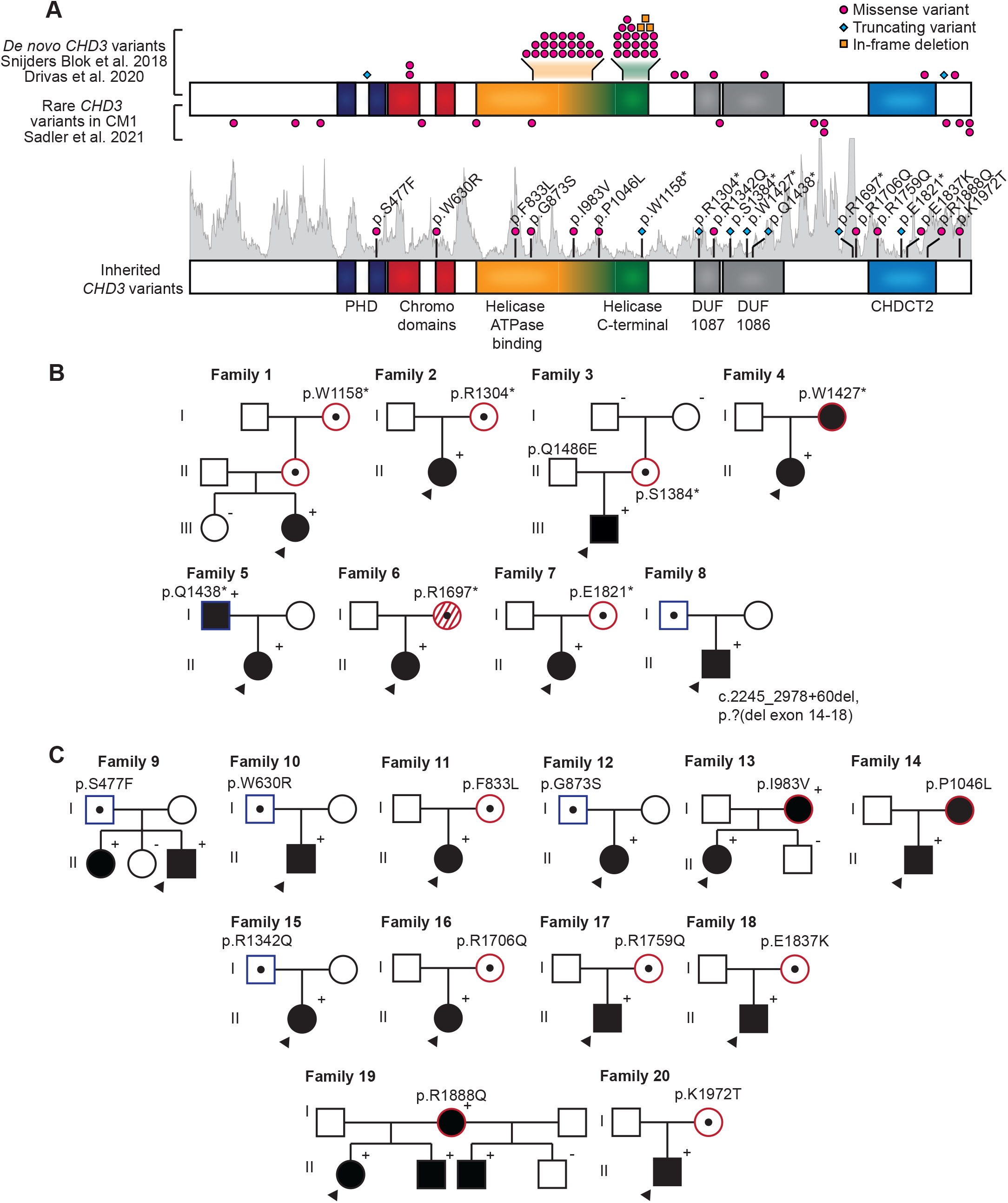
Twenty families with inherited *CHD3* variants. **A**) Schematic representation of the CHD3 protein (NM_001005273.2/NP_001005273.2), including functional domains, with PTVs labeled as cyan diamonds, in-frame deletions as orange squares, and missense variants as magenta circles. The intolerance landscape visualized using MetaDome ^70^ and computed based on single-nucleotide variants in the GnomAD database showing per amino acid position the missense over synonymous ratio, is shaded in gray. The top schematic shows the cases with *de novo CHD3* variants identified in NDD reported in ^12^ and ^13^ and rare variants associated with Chiari I malformations (CM1) reported in ^18^. The bottom schematic presents cases with inherited *CHD3* variants described in this study. **B-C**) Pedigrees of families identified with inherited *CHD3* variants, with in (B) families with predicted LoF variants and in (C) families with missense variants. The arrow head indicates the proband, filled symbols represent affected individuals (defined as individuals with developmental delay and/or intellectual disability), open symbols with a central dot represent confirmed carriers without developmental delay/intellectual disability, ‘+’ is used for a confirmed familial *CHD3* variant and ‘-’ for confirmed non-carriers. Symbols with red contours represent female carriers, symbols with blue contours represent male carriers. Dashed symbol for family 6 represents mosaic state of the variant in the mother. In pedigrees, only genetically tested siblings of the proband are shown.

**Figure 2.**
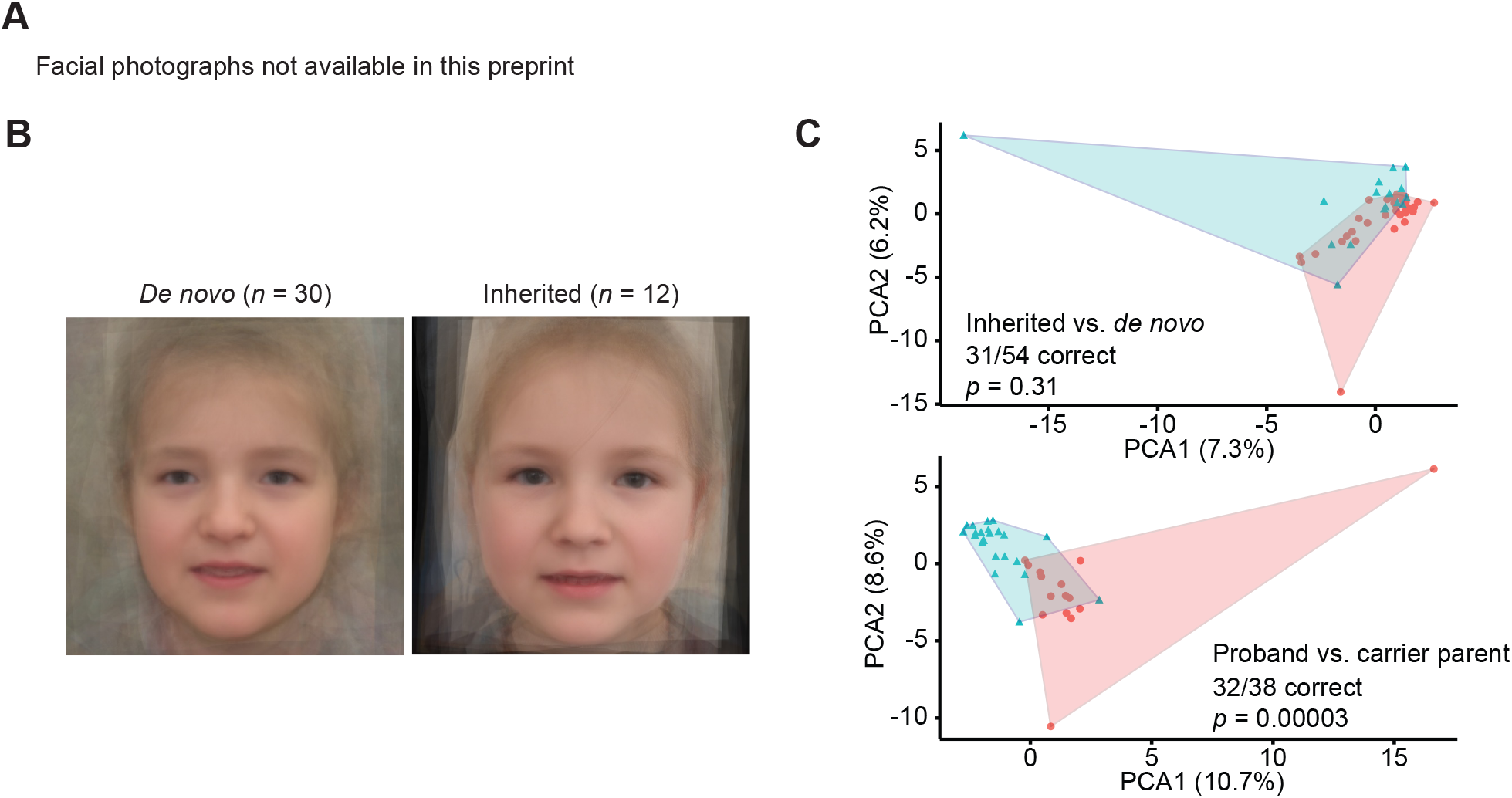
Facial features and clinical evaluation of individuals with inherited *CHD3* variants. **A**) Facial photographs of individuals with inherited *CHD3* variants. Individuals demonstrate features also observed in individuals with *de novo CHD3* variants, including a squared appearance of the face, prominent forehead, widely spaced eyes, thin upper lip, pointed chin and deep-set eyes. These characteristics are also present in carrier parents. As observed previously, facial gestalt changes with age ^13^. For example, a prominent nose is especially seen in adult individuals. For childhood pictures of carrier parents, see Figure S2. **B**) Computational average of facial photographs of 30 individuals with *de novo CHD3* variants (left) and 12 probands with inherited *CHD3* variants (right). **C**) Partitioning Around Medoids analyses of clustered HPO-standardized clinical data from 35 individuals with *de novo CHD3* variants, 19 affected probands with an inherited variant, and 19 carrier parents. The analyses do not show a significant distinction between the clusters of probands with *de novo* and probands with inherited variants (upper graph; *p* = 0.31). There is, however, a significant difference between the clusters of affected probands with inherited variants and carrier parents (bottom graph; *p* = 0.00003).

**Table 1:** Summary of phenotypes seen in individuals with *CHD3* variants.

| **Table 1: Summary of phenotypes seen in individuals with *CHD3* variants** |  |  |  |
| --- | --- | --- | --- |
|  | Probands with<br>*de novo* variant<sup>1</sup> | Probands with<br>inherited variant | Carrier<br>parents |
| **Development** |  |  |  |
| Developmental delay | 100% (55/55) | 100% (20/20) | 18% (3/17) |
| Intellectual disability | 98% (46/47) | 79% (11/14) | 22% (4/18) |
| *borderline/borderline-mild* | 6% (3/47) | 14% (2/14) | 11% (2/18) |
| *mild/mild-moderate* | 30% (14/47) | 29% (4/14) | 6% (1/18) |
| *moderate/moderate-severe* | 36% (17/47) | 14% (2/14) | 0% (0/18) |
| *severe* | 23% (11/47) | 0% (0/14) | 0% (0/18) |
| *level unknown* | 2% (1/47) | 21% (3/14) | 6% (1/18) |
| Speech delay/disorder | 100% (53/53) | 100% (19/19) | 25% (4/16) |
| Autism or autism-like features | 35% (18/51) | 56% (10/18) | 19% (3/16) |
| **Neurology** |  |  |  |
| Hypotonia | 81% (39/48) | 89% (16/18) | 18% (2/11) |
| Macrocephaly | 53% (28/53) | 44% (8/18) | 53% (8/15) |
| CNS abnormalities | 50% (24/48) | 62% (8/13) | 25% (1/4) |
| Neonatal feeding problems | 31% (10/32) | 17% (3/18) | 7% (1/15) |
| **Facial dysmorphisms** |  |  |  |
| High/broad/prominent forehead | 85% (28/33) | 84% (16/19) | 56% (9/16) |
| Thin upper lip | 79% (15/19) | 58% (11/19) | 44% (7/16) |
| Widely spaced eyes | 69% (35/51) | 74% (14/19) | 25% (4/16) |
| Broad nasal bridge | 75% (15/20) | 79% (15/19) | 25% (4/16) |
| Full cheeks | 58% (11/19) | 68% (13/19) | 7% (1/15) |
| Pointed chin | 60% (12/20) | 50% (9/18) | 38% (6/16) |
| Deep-set eyes | 55% (11/20) | 44% (8/18) | 47% (7/15) |
| **Other** |  |  |  |
| Joint laxity (generalized and/or local) | 36% (18/50) | 42% (8/19) | 31% (4/13) |
| Vision problems | 72% (38/53) | 26% (5/19) | 57% (8/14) |
| Male genital abnormalities | 32% (8/25) | 25% (2/8) | 0% (0/4) |
| Hernia (umbilical, inguinal, hiatal) | 13% (6/48) | 11% (2/19) | 0% (0/13) |
| <sup>1</sup>Combined cases from Snijders Blok et al. 2018 and Drivas et al. 2020 (confirmed *de novo* only) |  |  |  |
| CNS: central nervous system |  |  |  |

All of the carrier parents for whom phenotypic information was available about at least development and dysmorphisms (n = 6) had at least one feature of SNIBCPS (Table S2), although in five parents (31%) this was limited to only one (family 1 and 10) or two phenotypic features (family 9, 15 and 20; Table S2). Whereas the majority of carrier parents (15/16, 94%) presented with a single (e.g. prominent forehead or deep-set eyes) or several facial features known in SNIBCPS and 53% had macrocephaly (8/15) (Table 1 and S2), the parents had either mild/borderline intellectual disability (n = 4, 22%) or no history of intellectual disability (n = 14) (Table 1, S1 and S2; Supplemental Notes 1). Taken together, these observations suggest a combination of both variable expressivity and reduced penetrance for these rare genetic variations in *CHD3*.

We more objectively compared the phenotypes of probands with *de novo* and inherited *CHD3* variants based on Human Phenotype Ontology (HPO) terminology ^15^, using a Partitioning Around Medoids clustering algorithm ^16^. While this computational analysis did not identify a phenotypic difference between probands with *de novo* and inherited variants (31 of 54 individuals clustered correctly, *p* = 0.31; Figure 2C and S4), it confirmed a phenotypic difference between probands with inherited *CHD3* variants and their carrier parents (32 of 38 individuals clustered correctly, *p* = 0.00003; Figure 2C and S4). It is important to note that, although younger generations seem more severely affected than previous generations this may be due to ascertainment bias ^17^. Our finding of variable expressivity of inherited *CHD3* variants is consistent with the lack of a correlation between mutation location and phenotype ^13^, and the variable severity of phenotypes already described for (recurrent) *de novo CHD3* variants ^12; 13^. Additional evidence for variable expressivity for *CHD3* variation is provided by the recently identified association of 19 rare *CHD3* missense variants with Chiari I malformations in individuals without features of SNIBCPS ^18^.

We noticed that the majority of variants in our cohort were maternally inherited (14/20, 70%, *p* = 0.0577; Figure 1B, 1C and S5A). For single nucleotide variants with a LoF effect, 6/7 (86%, *p* = 0.0625) variants were maternally inherited (Figure 1B and S5A). Notably, the only father transmitting a LoF single nucleotide variant was affected (mild intellectual disability). This observation could hint at a female-protective effect for genetic variation in *CHD3*. Previous studies have repeatedly demonstrated a male bias in NDDs, a higher pathogenic variant burden in females and a maternal transmission bias in rare inherited variants ^6; 19-26^, suggesting that female gender protects against genetic variation in disease. This phenomenon might contribute to the variable expressivity observed for the inherited *CHD3* variants. However, we did not observe a sex-bias in the affected probands (12/20 female, *p* > 0.9999), or more severe intellectual disability in male compared to female *de novo* or inherited cases ^12; 13^. To further explore the hypothesis of a female protective effect at population level, we analyzed all *CHD3* LoF variants in gnomAD (n = 15 in 198,800 individuals) and found that females had a significantly higher carrier rate than males (12/15, *p* = 0.0173; Figure S5B).

Few cases with SNIBCPS have been described with confirmed *de novo CHD3* PTVs (4/55, 7.3% cases) ^12; 13^ including one which is predicted to escape nonsense-mediated decay (NMD; NP_001005273.1:p.Phe1935GlufsTer108). However, in our study we identified seven families with inherited single nucleotide PTVs and one with an intragenic deletion with a predicted LoF effect (8/20, 40%; Figure 1A). None of the inherited PTVs were predicted to escape NMD. We functionally confirmed this in family 1 (Figure 3A), for which we treated lymphoblastoid cell lines from the proband (individual III-2), carrier mother (II-2) and grandmother (I-2) and non-carrier healthy sibling of the proband (III-1) with cycloheximide to inhibit NMD, followed by direct amplification and Sanger sequencing of the *CHD3* transcript. We found that treatment with cycloheximide increased the expression of mutant allele, showing that the NM_001005273.2:c.3473G>A variant was targeted by NMD in all samples, as expected (Figure 3B).

**Figure 3.**
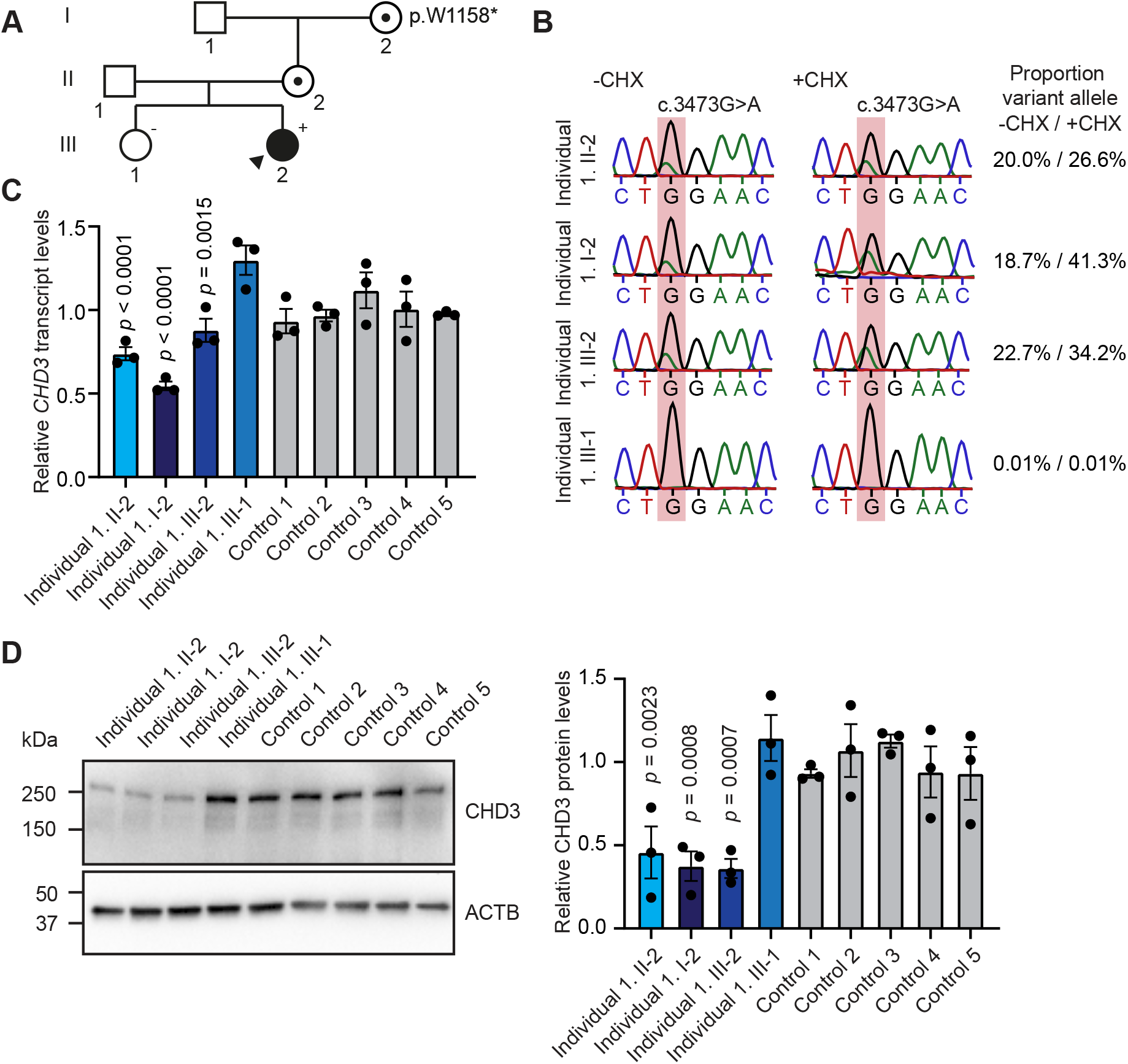
Functional consequences of the CHD3 p.W1158* PTV in subject-derived lymphoblastoid cell lines. **A**) Pedigree of family identified with an inherited *CHD3* c.3473G>A/p.W1158* variant **B**) Sanger sequencing chromatographs of EBV-immortalized lymphoblastoid cell lines derived from members of family 1. Individuals I-2, II-2 and III-2 carried the c.3473G>A/p.W1158* variant and individual III-1 was a healthy non-carrier sibling. Cells were treated with (+CHX) or without cycloheximide (-CHX) to test for NMD. The mutated position is shaded in red. The transcript carrying the variant allele is present at lower levels than the wild-type allele, and increases after CHX treatment (proportion variant allele calculated as: peak area variant allele / (peak area variant + wildtype allele), showing that this variant is targeted by NMD. **C**) qPCR of EBV-immortalized lymphoblastoid cell lines of family 1 (shades of blue) and five unrelated controls (gray) for *CHD3* transcript levels (NM_001005273.2). Values are normalized to expression of *PPIA* and *TBP* and shown relative to unrelated controls. Bars represent the mean with individual data points plotted (*n* = 3; *p*-values compared to individual III-1 (healthy non-carrier sibling), one-way ANOVA and post hoc Bonferroni test). **D**) Left, a representative immunoblot of protein lysates prepared from lymphoblastoid cell lines for CHD3 (expected molecular weight: ∼227 kDa). The blot was probed for ACTB, to ensure equal protein loading. Right, a graph showing the quantification of immunoblots with bars presenting the mean and individual data points plotted (*n* = 3; *p*-values compared to individual III-1 (healthy non-carrier sibling), one-way ANOVA and post hoc Bonferroni test). Controls are shaded in gray and samples from family 1 are shaded in blue. **C-D**) The cell lines carrying c.3473G>A/p.W1158* show lower *CHD3* transcript/protein levels compared to the control samples.

An explanation for variable expressivity of PTVs could be compensation of expression by the wildtype allele to maintain normal expression levels ^27^. To test if such compensation plays a role in variable expressivity of *CHD3* PTVs, we evaluated the expression of the *CHD3* variant in family 1 (c.3473G>A, p.W1158*) on a transcript and protein level. We found that this variant resulted in lower levels of *CHD3* transcript and CHD3 protein levels, when compared to lymphoblastoid cells from the non-carrier healthy sibling (Figure 3C and 3D). These findings confirm the LoF effect of the stop-gain variant in this family, and makes compensation by the wildtype allele as an underlying mechanism for the milder phenotype in the carrier mother and grandmother unlikely. It remains, however, to be determined, whether such LoF variance can have a tissue-specific, temporal expression specific, and/or transcript specific effect. It is unclear whether results from blood-derived cells can be extrapolated to neuronal cell types, which would be more relevant considering the NDD phenotypes in our cohort, especially given that neuron-specific alternative splicing has previously been described for *CHD3* ^28^. Other explanations for the clinical variable expressivity of inherited *CHD3* variants include the presence of a second-hit on the other allele by either rare or common variation, a genome wide higher mutational burden of high-penetrant variants, or common variants in promoter/enhancers regions or in other genes, inherited from the non-carrier parent ^9; 29; 30^. Such a compound inheritance mechanism, has, for example, been described for thrombocytopenia with absent radii (TAR) syndrome, where the inheritance of a rare null allele together with one of two low-frequency SNPs in regulatory regions causes disease ^31^. In four probands with an inherited *CHD3* variant a copy number variant (CNV) was also reported, including one 22q11.2 duplication, which has been described with highly variable features (MIM # 608363) and three CNVs of unknown significance (Table S1). Proband 7 had other (likely) pathogenic variants contributing to the phenotype (Table S1; Supplemental Notes 1). A comparison with the prevalence of additional genetics finding in individuals with *de novo CHD3* variants could not be made due to lack of reporting on additional genetic findings ^12; 13^.

In addition to the seven inherited *CHD3* single nucleotide PTVs and the intragenic deletion, we identified 12 families with inherited missense variants. One of the identified inherited missense variants also present in an unaffected carrier parent was identical to a variant previously reported as a *de novo* variant in an individual with SNIBCPS (p.R1342Q; individual 32 in ^12^). Based on the phenotypes observed in the probands with inherited *CHD3* missense variants, the conservation of affected positions (Figure S1), and *in silico* predictions of pathogenicity (Figure S6; Table S1), we considered these inherited *CHD3* missense variants as likely pathogenic with variable expressivity in the parents. Clinically, probands carrying a *CHD3* missense variant did not seem to be more severely affected than individuals with PTVs (Table S3). The individuals with *de novo* missense variants published to date were mostly (although not entirely) localized to the ATPase-Helicase domain ^12; 13^. No clustering to the ATPase-Helicase domain or elsewhere was observed among the inherited missense variants of our cohort (Figure 1A). It has been speculated that the *de novo* missense variants clustering in the ATPase-Helicase domain are unlikely to lead to a sole LoF effect ^12^, and may potentially act in a dominant-negative way. The identification of eight families with an inherited LoF variant and the lack of clustering of the inherited missense variants may suggest a LoF effect as the main mechanism for inherited cases, which may underlie the variable expressivity. However, our cell-based analyses of chromatin binding (for p.S477F) and GATAD2B-binding (for p.R1342Q, p.E1837K and p.Q1888R) did not find evidence of LoF for the protein functions that we tested (Figure S7). This does not exclude that these variants have an effect on other biological functions of CHD3. Based on 3D-protein modeling, the prior published *de novo* missense variants within the ATPase-Helicase domain localize more closely to the ATP-binding site than the inherited missense variants of our cohort (Supplemental Notes 2). Interestingly, the p.I983V (family 13) variant was found to be closer to published *de novo* variants (Supplemental Notes 2) and the carrier parent with this missense variant did have a neurodevelopmental phenotype which was more pronounced than in other carrier parents (Figure 1C and 2; Table S2; Supplemental Notes 1).

The presence of rare, likely pathogenic *CHD3* variants in healthy individuals prompted us to study possible effects of variation in this gene at a population level, using data from the UK Biobank resource ^32-37^. For a detailed description of these analyses, see Supplemental Notes 3. We found no associations between rare missense variation at mutation-intolerant locations in *CHD3* (minor allele frequency ≤ 1%, located in functional domains, damaging in PolyPhen or SIFT and with a CADD-PHRED score > 25) and fluid intelligence (N = 77,998), educational qualification (N = 120,596) or intracranial volume (N = 18,254). However, we identified a group difference in intracranial volume between rare *CHD3* putative LoF variant carriers and non-carriers, with carriers having an increased volume compared to controls (n = 4, *t* = 2.37, *p* = 0.018). This seems consistent with the observation of macrocephaly in 44-53% of probands with a (likely) pathogenic *CHD3* variant and in 53% of carrier parents (Table 1), and the link of rare *CHD* variants with abnormal brain growth ^18^. To test possible relationships between *CHD3* common genetic variation and head circumference and/or intracranial volume, we performed gene-level analyses using previously published SNP-wise association summary statistics for these traits ^38; 39^, but no tests survived multiple testing correction (Supplemental Notes 3).

With the identification and characterization of inherited *CHD3* variants with variable expressivity in twenty families, we showed that, in addition to highly penetrant *de novo* variants, rare predicted likely pathogenic inherited variants in *CHD3* should be considered as possibly pathogenic depending on mutation characteristics in cases with phenotypic concordance to SNIBCPS. Interestingly, variable penetrance and expressivity has been noted in numerous families with another dominant NDD, KBG syndrome, caused by LoF variants in *ANKRD11* (MIM #148050) ^40; 41^. So this phenomenon is likely more common for dominant NDDs, with important implications for clinical genetic counseling, in the context of recurrence risk, prenatal diagnostics, prognosis and variant interpretation.

Clinically, we recommend that it can be helpful to evaluate the parents of children with *CHD3* variants for subtle SNIBCPS features. In particular macrocephaly and facial dysmorphisms including a prominent forehead and pointed chin could be recognized in a substantial number of carrier parents (53% and 94% respectively; Figure 2A; Table S2). Taken together, our results illustrate the continuum of causality for NDDs with genetic origins ^17; 42^ and significantly underline the hypothesis that variable expressivity and reduced penetrance likely explain a large portion of as yet unexplained NDD cases. Overall, we show that even for genes already known to be implicated in a NDD inherited variation and variable expressivity can play a major role and are thus important to consider in genetic counseling.

## Data Availability

Supplementary Notes 1, Table S1, facial photographs (Fig. 2A and Fig. S2) and all datasets generated and analyzed during the current study are available from the corresponding author on request.

## Acknowledgements

We are extremely grateful to all families participating in this study. In addition, we wish to thank the members of the Cell culture facility, Department of Human Genetics, RadboudUMC, Nijmegen for culture of cell lines. This work was financially supported by the Dutch Research Council grant to TK (015.014.036 and 1160.18.320) and LELMV (015014066) and Netherlands Organization for Health Research and Development to TK (91718310) and LELMV (843002608, 846002003), by Donders Junior Researcher Grant 2019 to TK and LELMV and the Max Planck Society (JdH, DS, CF, SEF). Authors of this publication are members of the European Reference Network on Rare Congenital Malformations and Rare Intellectual Disability ERN-ITHACA [EU Framework Partnership Agreement ID: 3HP-HP-FPA ERN-01-2016/739516]. The aims of this study contribute to the Solve-RD project (CG, HGB, LELMV and TK) which has received funding from the European Union’s Horizon 2020 research and innovation programme under grant agreement No 779257. For families 10,13,14 and 19 sequencing and analysis were provided by the Broad Institute of MIT and Harvard Center for Mendelian Genomics (Broad CMG) and were funded by the National Human Genome Research Institute, the National Eye Institute, and the National Heart, Lung and Blood Institute grant UM1 HG008900 and in part by National Human Genome Research Institute grant R01 HG009141. For family 15 exome sequencing was performed in the framework of the German project “TRANSLATE NAMSE”, an initiative from the National Action League for People with Rare Diseases (Nationales Aktionsbündnis für Menschen mit Seltenen Erkrankungen, NAMSE) facilitating genetic diagnostics for individuals with suggested rare diseases. Part of this research has been conducted using the UK Biobank Resource under application number 16066, with Clyde Francks as the principal applicant. Our study made use of imaging-derived phenotypes generated by an image-processing pipeline developed and run on behalf of UK Biobank.

## Materials and Methods

### Individuals and consent

The cohort presented in this study was assembled from hospitals and laboratories across the Netherlands, Germany, United States of America, Slovenia, Australia and Canada. Informed consent for the use and publication of medical data and biological material was obtained from all patients or their legal representative by the involved clinician. Consent for publication of photographs was obtained separately. Genetic testing and research were performed in accordance with protocols approved by the local Institutional Review Boards.

### Next-generation-sequencing

*CHD3* variants in all probands were identified using whole exome sequencing (WES) or whole genome sequencing (WGS; family 4 and 12), with filtering as previously described ^3; 43-52^. Due to inheritance from seemingly healthy/mildly affected parents the *CHD3* variants were initially classified as variants of unknown significance. Inheritance of variants was confirmed either as part of trio WES or using targeted Sanger sequencing after identification in singleton exon analysis. Similarly, if applicable, other family members were tested using targeted sequencing.

Pathogenicity of missense variants was further evaluated using CADD-PHRED v1.6 ^53^, PolyPhen-2 ^54^ and SIFT ^55^ scores. Allele frequencies of all variants in gnomAD were based on ENST00000330494.7 ^14^.

### Facial analysis

We established a 2D hybrid facial model which combines the analysis of the ‘Clinical Face Phenotype Space’ pipeline with the facial recognition system of the ‘OpenFace’ pipeline ^56; 57^. First, we generated a 468-dimensional feature vector of the facial features of 30 individuals with *de novo CHD3* variants. After extraction of the hybrid features for each of the individuals, we calculated whether the individuals with *de novo CHD3* variants cluster together when compared to a group of matched controls based on the nearest neighbor principle (Euclidean distance) – these matched controls were individuals with ID and are age-, ethnicity- and gender matched. The Mann-Withney U test was used to determine whether the clustering of individuals with *de novo CHD3* variants was significantly higher than expected based on random chance. A *p*-value smaller than 0.05 was considered significant.

Furthermore, a classifier was built using a logistic regression model trained on the 468-dimensional feature vector of the 30 individuals. The performance was evaluated performing leave-one-out cross validation and the classifier was shown to have a sensitivity of 0.91, a specificity of 0.83 and an overall area under the ROC-curve of 0.91. Finally, using the trained classifier, we determined for each inherited case whether that individual clusters within the *de novo CHD3* group or the control group (Figure S3).

### Construction of composite face

For 12 individuals with an inherited *CHD3* variant and 30 with a *de novo CHD3* variant, facial 2D-photographs were available for generating a composite face. As previously described, average faces were generated while allowing for asymmetry preservation and equal representation by individuals ^58^.

### Human Phenotype Ontology (HPO)-based phenotype clustering analysis

HPO-based clustering analysis was performed as described elsewhere ^59^. We included 35 individuals with *de novo CHD3* variants ^12^, 19 of 20 probands with an inherited *CHD3* variant, and 19 of 20 carrier parents in the analysis: the proband and carrier mother of family 6 were excluded because no clinical data were available, and the mother is mosaic for the *CHD3* variant (∼37%). The Wang score (a measure of semantic similarity) between all terms was calculated using the HPO Sim package ^60; 61^. The terms were divided in groups, based on the similarity score: a new feature – the sum of the terms in the group - was created as a replacement for the terms in that specific group (Figure S4A; Table S4). HPO terms that could not be added to a group feature were added as a separate term. To quantify and visualize possible differences in our cohort, we used Partitioning Around Medoids (PAM) clustering on these grouped features. We compared probands with a *de novo* and inherited variant and probands with inherited variants and their carrier parents in a second analysis. To assess statistical significance, a permutations test (*n* = 100,000) was used with relabeling based on variant types, while keeping the original distribution of variant types into account.

### Three-dimensional protein modeling

We modeled the protein structure of the ATPase-Helicase domain of CHD3 in interaction with the DNA using the homology modeling script in the WHAT IF ^62^ & YASARA ^63^ Twinset with standard parameters. As a template, we used PDB file 6RYR which contains the human Nucleosome-CHD4 complex structure of a single copy of CHD4 ^64^. The PHD2 variant (p.S477F) was modeled in the PHD2 domain of CHD4 (PDB 2L75, 89% sequence identity with CHD3) ^65^.

### DNA expression constructs and site-directed mutagenesis

The cloning of *CHD3* (NM_001005273.2/ENST00000330494.7) has been described previously ^12^. The coding DNA sequence of *GATAD2B* (NM_020699.3/ENST00000368655.4) and a C-terminal region of CHD3-encoding residues 1246-1944 (NM_001005273.2) were amplified using primers listed in Table S6. Variants in full-length CHD3 or the C-terminal CHD3 construct were generated using the QuikChange Lightning Site-Directed Mutagenesis Kit (Agilent). The primers used for site-directed mutagenesis are listed in Table S7. cDNAs were subcloned using BamHI/HpaI (full-length CHD3), BamHI/XbaI (GATAD2B) or HindIII/BamHI (C-terminal CHD3 construct) into pYFP, pHisV5 and pRluc, created by modification of the pEGFP-C2 vector (Clontech) as described before ^66^. All constructs were verified by Sanger sequencing.

### Cell culture

Lymphoblastoid cell lines (LCLs) were established by Epstein-Barr virus transformation of peripheral lymphocytes from blood samples collected in heparin tubes, and maintained in RPMI medium (Sigma) supplemented with 15% fetal bovine serum and 5% HEPES (both Invitrogen). HEK293T/17 cells (CRL-11268, ATCC) were grown in DMEM supplemented with 10% fetal bovine serum and 1x penicillin-streptomycin (all Invitrogen) at 37°C with 5% CO2. Transfections were performed using GeneJuice (Millipore) following the manufacturer’s protocol.

### Testing for nonsense mediated decay of truncating variants

LCLs of members of family 1 and controls were grown overnight with 100 µg/ml cycloheximide (Sigma) to block NMD. After treatment, cell pellets were collected, and RNA and protein were extracted using the RNeasy Mini Kit (Qiagen) or with 1x RIPA buffer supplemented with 1% PMSF and 1x PIC, respectively. RT-PCR was performed using SuperScript III Reverse Transcriptase (ThermoFisher) with random primers, and regions of interest were amplified from cDNA using primers listed in Table S5. Sanger trace peak sizes of the wildtype and variant allele were measured using the ‘Area’ option in ImageJ and proportion of the variant allele was calculated: peak area variant allele / (peak area variant + wildtype allele).

### Direct fluorescent imaging

HEK293T/17 cells were grown on coverslips coated with poly-D-lysine (Sigma). Forty-eight hours after transfection with the YFP-tagged C-terminal CHD3 construct and HisV5-tagged GATAD2B, cells were fixed with 4% paraformaldehyde (PFA, Electron Microscopy Sciences). Nuclei were stained with Hoechst 33342 (Invitrogen). Fluorescence images were acquired with a Zeiss LSM880 confocal microscope and Airyscan unit using ZEN Image Software (Zeiss).

### FRAP assays

HEK293T/17 cells were transfected in clear-bottomed black 96-well plates with YFP-tagged full-length CHD3 or p.S477F. After 48 h, medium was replaced with phenol red-free DMEM supplemented with 10% fetal bovine serum (both Invitrogen), and cells were moved to a temperature-controlled incubation chamber at 37°C. Fluorescent recordings were acquired using a Zeiss LSM880 and Zen Black Image Software, with an alpha Plan-Apochromat 100x/1.46 Oil DIC M27 objective (Zeiss). FRAP experiments were performed by photobleaching an area of 0.98 μm x 0.98 μm within a single nucleus with 488-nm light at 100% laser power for three iterations with a pixel dwell time of 32.97 μs, followed by collection of times series of 150 images with a 2.5 zoom factor and an optical section thickness of 1.4 μm (2.0 Airy units). Individual recovery curves were background subtracted and normalized to the pre-bleach values, and mean recovery curves were calculated using EasyFRAP software ^67^. Curve fitting was done with the FrapBot application using direct normalization and a single component exponential model, to calculate the half-time and maximum recovery ^68^.

### Immunoblotting

Whole-cell lysates were collected in 1x RIPA buffer (ThermoFisher) supplemented with 1x PIC (Roche) and 1% PMSF (Sigma). Cells were lysed for 20 min at 4 °C followed by centrifugation for 20 min at 12,000 rpm. Samples were loaded on 4–15% Mini-PROTEAN TGX Precast Gels (Bio-Rad) and transferred onto polyvinylidene fluoride membranes. Membranes were blocked in 5% milk for 1 h at room temperature and then probed with rabbit-anti-CHD3 antibody (1:1000; Abcam, ab109195) or mouse-anti-GFP (1:8000; Clontech, 632380) overnight at 4°C. Next, membranes were incubated with HRP-conjugated goat-anti-rabbit or goat-anti-mouse antibody (1:10,000; Jackson ImmunoResearch) for 1.5 h at room temperature. Bands were visualized with the SuperSignal West Femto Maximum Sensitivity Substrate Reagent Kit (CHD3; ThermoFisher) or the Novex ECL Chemiluminescent Substrate Reagent Kit (YFP-fusion proteins; Invitrogen) using a ChemiDoc XRS + System (Bio-Rad).

### Co-immunoprecipitation

HEK293T/17 cells were transfected with the YFP-tagged C-terminal region of CHD3 and Rluc-tagged GATAD2B. After 48h, whole-cell lysates were collected in Pierce IP Lysis Buffer (25 mM Tris-HCl pH 7.4, 150 mM NaCl, 1 mM EDTA, 1% NP-40 and 5% glycerol; ThermoFisher) supplemented with 1x PIC (Roche) and 1% PMSF (Sigma). Cells were lysed for 20 min at 4°C followed by centrifugation for 20 min at 12,000 rpm. YFP-fusion proteins were immobilized on GFP-trap magnetic agarose beads (Chromotek) overnight at 4°C. Deactivated beads (Chromotek) were used as a negative control. The elutions and 5% of the input were resolved on 4–15% Mini-PROTEAN TGX Precast Gels (Bio-Rad) and transferred onto polyvinylidene fluoride membranes. Membranes were blocked in 5% milk for 1 h at room temperature and then probed with rabbit-anti-Rluc antibody (1:2000; GeneTex) overnight at 4°C. Next, membranes were incubated with HRP-conjugated goat-anti-rabbit antibody (1:10,000; Jackson ImmunoResearch) for 1.5 h at room temperature. Bands were visualized with the SuperSignal West Femto Maximum Sensitivity Substrate Reagent Kit (ThermoFisher) using a ChemiDoc XRS + System (Bio-Rad).

### Population-based analysis of the association of CHD3 variation with intelligence, educational qualification and intracranial volume/head circumference

Using WES data of 200,000 individuals from the UKB Exome Sequencing Consortium (^33; 34^, and https://www.ukbiobank.ac.uk/media/cfulxh52/uk-biobank-exome-release-faq_v9-december-2020.pdf) we studied the association of *CHD3* missense and putative LoF variants with ‘Fluid intelligence score’ (data field ID 3533), ‘Qualifications’ (data field ID 6138) and ‘Volume of EstimatedTotalIntraCranial’ (data field ID 7054). Additionally, we used genome-wide association meta-analysis summary statistics of head circumference (N ≤ 18,881), and HC combined with intracranial volume (N ≤ 45,458) in child- and adulthood ^38^, and infant head circumference (N ≤ 10,768) ^39^ to calculate gene-level p-values reflecting the common variant associations of *CHD3* with these traits using MAGMA ^69^. For detailed description of the methods, see Supplemental Notes 3.

