## Supplemental information for "Inherited variants in *CHD3* demonstrate variable expressivity in Snijders Blok-Campeau syndrome"

p.S477F

```
TR|F6PUX9|F6PUX9_XENTR CCDACVSSYHIHCLNPPLPDIPHGEWLCPRCTCPQLKGKVQKILHWRWGVPPEGVPLPQP 550
SP|Q12873|CHD3_HUMAN   CCDACISSYHIHCLNPPLPDIPNGEWLCPRCTCPVLKGRVQKILHWRWGEPPVAVPAPQQ 529
TR|B1AR17|B1AR17_MOUSE CCDACISSYHIHCLNPPLPDIPNGEWLCPRCTCPVLKGRVQKILHWRWGEPPVAVPAPQQ 581
TR|F1LPP8|F1LPP8_RAT   CCDACISSYHIHCLNPPLPDIPNGEWLCPRCTCPVLKGRVQKILHWRWGEPPVAVPAPQQ 361
                       *****:****************:*********** ***:********** ** .** **
```

pW630R

```
TR|F6PUX9|F6PUX9_XENTR PPPFDYGSGEDEGKSEK--SKDAEYSDLEERFYRYGIKPEWMSIQRIINHSLDKRGNYHY 668
SP|Q12873|CHD3_HUMAN   PPPLDYGSGEDDGKSDKRKVKDPHYAEMEEKYYRFGIKPEWMTVHRIINHSVDKKGNYHY 649
TR|B1AR17|B1AR17_MOUSE PPPLDYGSGEDDGKSDKRKVKDPHYAEMEEKYYRFGIKPEWMTVHRIINHSMDKKGNYHY 701
TR|F1LPP8|F1LPP8_RAT   PPPLDYGSGEDDGKSDKRKVKDPHYAEMEEKYYRFGIKPEWMTVHRIINHSMDKKGNYHY 481
                       ***:*******:***:*   ** .*:::**::**:*******:::******:**:*****
```

p.F833L p.G873S

```
TR|F6PUX9|F6PUX9_XENTR EFSYQDNVMKGGKKAFKMKAQAQVKFHVLLTSYELVTIDQAALGSIRWACLVVDEAHRLK 908
SP|Q12873|CHD3_HUMAN   EFSFEDNAIKGGKKAFKMKREAQVKFHVLLTSYELITIDQAALGSIRWACLVVDEAHRLK 889
TR|B1AR17|B1AR17_MOUSE EFSFEDNAIKGGKKAFKMKREAQVKFHVLLTSYELITIDQAALGSIRWACLVVDEAHRLK 941
TR|F1LPP8|F1LPP8_RAT   EFSFEDNAIKGGKKAFKMKREAQVKFHVLLTSYELITIDQAALGSIRWACLVVDEAHRLK 721
                       ***::**.:********** :**************:************************
```

p.I983V

```
TR|F6PUX9|F6PUX9_XENTR EDQIKKLHDLLGPHLLRRMKADVFKNMPAKTELIVRVELSPMQKKYYKFILTRNFEALNS 1028
SP|Q12873|CHD3_HUMAN   EDQIKKLHDLLGPHMLRRLKADVFKNMPAKTELIVRVELSPMQKKYYKYILTRNFEALNS 1009
TR|B1AR17|B1AR17_MOUSE EDQIKKLHDLLGPHMLRRLKADVFKNMPAKTELIVRVELSPMQKKYYKYILTRNFEALNS 1061
TR|F1LPP8|F1LPP8_RAT   EDQIKKLHDLLGPHMLRRLKADVFKNMPAKTELIVRVELSPMQKKYYKYILTRNFEALNS 841
                       **************:***:*****************************:***********
```

p.P1046L

```
TR|F6PUX9|F6PUX9_XENTR RGGGNQVSLLNIMMDLKKCCNHPYLFPAASLESPKLPSGAYEGGSLVKASGKLLLLHKML 1088
SP|Q12873|CHD3_HUMAN   RGGGNQVSLLNIMMDLKKCCNHPYLFPVAAMESPKLPSGAYEGGALIKSSGKLMLLQKML 1069
TR|B1AR17|B1AR17_MOUSE RGGGNQVSLLNIMMDLKKCCNHPYLFPVAAMESPKLPSGAYEGGALIKSSGKLLLLQKML 1121
TR|F1LPP8|F1LPP8_RAT   RGGGNQVSLLNIMMDLKKCCNHPYLFPVAAMESPKLPSGAYEGGALIKSSGKLLLLQKML 901
                       ***************************.*::*************:*:*:****:**:***
```

p.R1342Q

```
TR|F6PUX9|F6PUX9_XENTR QEENVDPDYWEKLLRHHYEQQQEDLARNLGKGKRVRKQVNYNDAAQEDQDNQSEYSVGSE 1388
SP|Q12873|CHD3_HUMAN   QEENVDPDYWEKLLRHHYEQQQEDLARNLGKGKRVRKQVNYNDAAQEDQDNQSEYSVGSE 1368
TR|B1AR17|B1AR17_MOUSE QEENVDPDYWEKLLRHHYEQQQEDLARNLGKGKRVRKQVNYNDAAQEDQDNQSEYSVGSE 1420
TR|F1LPP8|F1LPP8_RAT   QEENVDPDYWEKLLRHHYEQQQEDLARNLGKGKRVRKQVNYNDAAQEDQDNQSEYSVGSE 1200
                       ************************************************************
```

p.R1706Q

```
TR|F6PUX9|F6PUX9_XENTR PRFMFNIADGGFTELHTLWQNEERAAICSGRLNDIWHRRHDYWLLAGIVVHGYARWQDIQ 1807
SP|Q12873|CHD3_HUMAN   PRFMFNIADGGFTELHTLWQNEERAAISSGKLNEIWHRRHDYWLLAGIVLHGYARWQDIQ 1764
TR|B1AR17|B1AR17_MOUSE PRFMFNIADGGFTELHTLWQNEERAAISSGKLNEIWHRRHDYWLLAGIVLHGYARWQDIQ 1817
TR|F1LPP8|F1LPP8_RAT   PRFMFNIADGGFTELHTLWQNEERAAISSGKLNEIWHRRHDYWLLAGIVLHGYARWQDIQ 1597
                       ***************************.**:**:***************:**********
```

p.R1759Q

```
TR|F6PUX9|F6PUX9_XENTR PRFMFNIADGGFTELHTLWQNEERAAICSGRLNDIWHRRHDYWLLAGIVVHGYARWQDIQ 1807
SP|Q12873|CHD3_HUMAN   PRFMFNIADGGFTELHTLWQNEERAAISSGKLNEIWHRRHDYWLLAGIVLHGYARWQDIQ 1764
TR|B1AR17|B1AR17_MOUSE PRFMFNIADGGFTELHTLWQNEERAAISSGKLNEIWHRRHDYWLLAGIVLHGYARWQDIQ 1817
TR|F1LPP8|F1LPP8_RAT   PRFMFNIADGGFTELHTLWQNEERAAISSGKLNEIWHRRHDYWLLAGIVLHGYARWQDIQ 1597
                       ***************************.**:**:***************:**********
```

p.E1837K

```
TR|F6PUX9|F6PUX9_XENTR PAMALNARFSEVECLAESHQHLSKESLAGNKPANAVLHKVLNQLEELLSDMKADVTRLPA 1927
SP|Q12873|CHD3_HUMAN   PAMALHARFAEAECLAESHQHLSKESLAGNKPANAVLHKVLNQLEELLSDMKADVTRLPA 1884
TR|B1AR17|B1AR17_MOUSE PAMALHARFAEAECLAESHQHLSKESLAGNKPANAVLHKVLNQLEELLSDMKADVTRLPA 1937
TR|F1LPP8|F1LPP8_RAT   PAMALHARFAEAECLAESHQHLSKESLAGNKPANAVLHKVLNQLEELLSDMKADVTRLPA 1717
                       *****:***:*.************************************************
```

p.R1888Q

```
TR|F6PUX9|F6PUX9_XENTR SLARIPPIAARLQMSERSILSRLASKGSESTAPPVFPPGPYTTPPMFGATFPSNPQSA-A 1986
SP|Q12873|CHD3_HUMAN   TLSRIPPIAARLQMSERSILSRLASKGTEPHPTPAYPPGPYATPPGYGAAFSAAPVGALA 1944
TR|B1AR17|B1AR17_MOUSE TLSRIPPIAARLQMSERSILSRLASKGTEPHPTPAFPPGPYATPPGYGAAFSAAPVGALA 1997
TR|F1LPP8|F1LPP8_RAT   TLSRIPPIAARLQMSERSILSRLASKGTEPHPTPAFPPGPYATPPGYGAAFSAAPVGALA 1777
                       :*:************************:*    *.:*****:*** :**:* : * .* *
```

p.K1972T

```
TR|F6PUX9|F6PUX9_XENTR LTGTNYNQMPLGSFLSA-SNGPPVLVKKEREMDL--------LDRKESRGGEVICIDD 2035
SP|Q12873|CHD3_HUMAN   AAGANYSQMPAGSFITAATNGPPVLVKKEKEMVGALVSDG--LDRKEPRAGEVICIDD 2000
TR|B1AR17|B1AR17_MOUSE AAGANYSQMPAGSFITATTNGPPVLVKKEKEMVGALVSDGLGLDRKEPRAGEVICIDD 2055
TR|F1LPP8|F1LPP8_RAT   AAGANYSQMPAGSFITATTNGPPVLVKKEKEMVGALVSDGLGLDRKEPRAGEVICIDD 1835
                        :*:**.*** ***::* :**********:**          ***** *.********
```

**Figure S1. Amino acid sequence alignments of the CHD3 protein.** Amino acid sequences of the human CHD3 protein (Q01826, UniProt) aligned to the Xenopus tropicalis (F6PUX9), mouse (B1AR17) and rat (F1LPP8) sequences. Alignment was performed with Clustal Omega (1.2.4) with default settings using EMBL-EBI alignment tool. Missense variants described in this study are shaded in red.

Facial photographs not available in this preprint

**Figure S2. Childhood images of parents carrying a *CHD3* variant.** Facial photographs taken during childhood of parents found to carry a *CHD3* variant. Individuals 1. I-2, 1. II-2 and 16. I-2 are four years old in the picture, individual 11. I-2 is one year old. Individual 18. I-2, from left to right, as infant, toddler and child. Images show several features also seen in probands with *CHD3* variants including a squared face, pointed chin, deep-set eyes and a broad nasal bridge. These features become less prominent when these individuals get older (compare to Figure 2A).

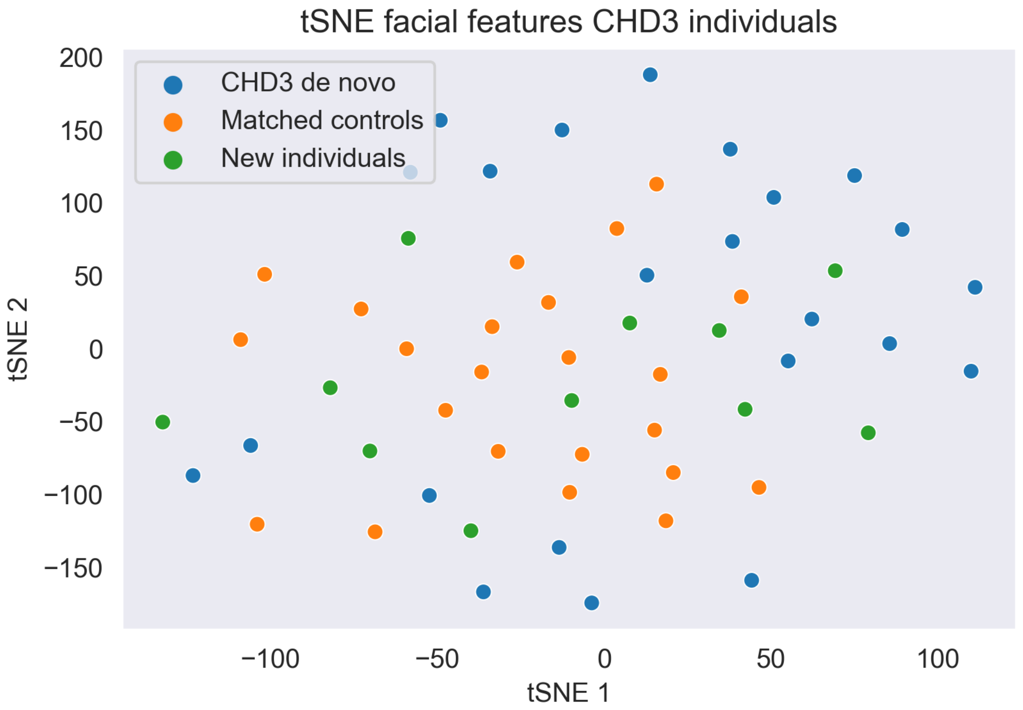


**Figure S3. t-SNE plot of the distribution of individuals with *de novo* and inherited *CHD3* variants versus controls with intellectual disability.** t-distributed stochastic neighbor embeddings (t-SNE) plot that visualizes the distribution of the hybrid feature vectors for individuals with *de novo* ('CHD3 de novo'; blue) and inherited *CHD3* variants ('New individuals'; green), and intellectual disability (ID) controls ('Matched controls'; orange). Relative clustering of individuals with a *de novo* variant in *CHD3* suggests that this group of individuals with intellectual disability shows more similarity in facial features than is expected by chance (AROC: 0.91). The control group was matched to the *de novo CHD3* group for gender, ethnicity, and age. Individuals with an inherited *CHD3* variant were tested for clustering with either the *de novo CHD3* group, or the ID controls group, based on the nearest-neigbor principle (see Table S2 for results per individual).

**A**

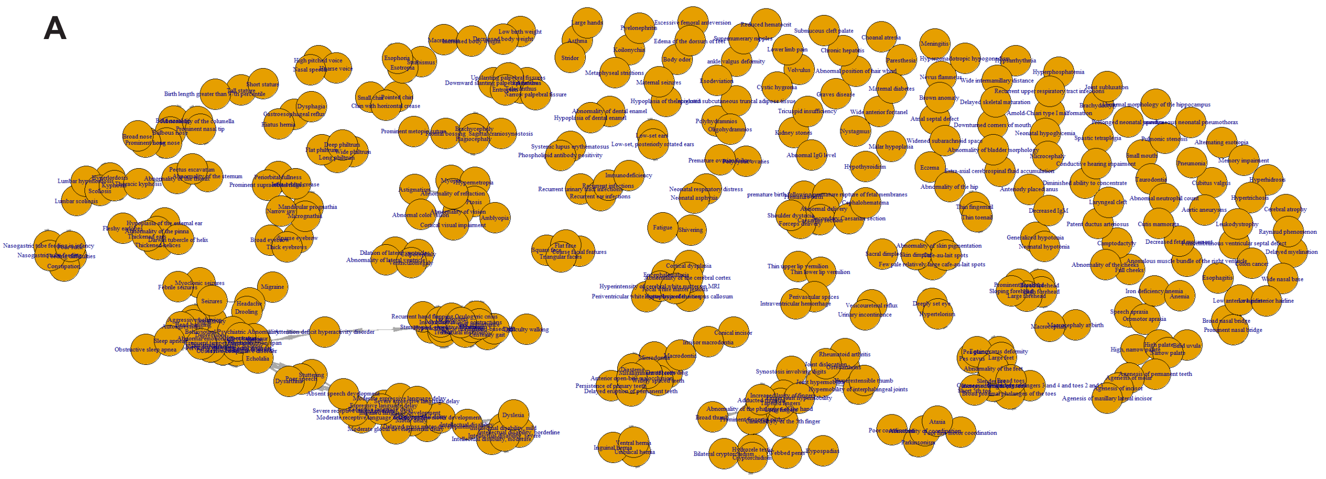

**B**

| Cluster | Labels_reversed | Index | CORRECT_CLASSIFICATIE |
|---|---|---|---|
| 1 | 0 | CHD3_dn_001 | FALSE |
| 0 | 0 | CHD3_dn_002 | TRUE |
| 1 | 0 | CHD3_dn_003 | FALSE |
| 0 | 0 | CHD3_dn_004 | TRUE |
| 0 | 0 | CHD3_dn_005 | TRUE |
| 1 | 0 | CHD3_dn_006 | FALSE |
| 0 | 0 | CHD3_dn_007 | TRUE |
| 0 | 0 | CHD3_dn_008 | TRUE |
| 0 | 0 | CHD3_dn_009 | TRUE |
| 0 | 0 | CHD3_dn_010 | TRUE |
| 0 | 0 | CHD3_dn_011 | TRUE |
| 1 | 0 | CHD3_dn_012 | FALSE |
| 0 | 0 | CHD3_dn_013 | TRUE |
| 0 | 0 | CHD3_dn_014 | TRUE |
| 0 | 0 | CHD3_dn_015 | TRUE |
| 0 | 0 | CHD3_dn_016 | TRUE |
| 0 | 0 | CHD3_dn_017 | TRUE |
| 0 | 0 | CHD3_dn_018 | TRUE |
| 1 | 0 | CHD3_dn_019 | FALSE |
| 0 | 0 | CHD3_dn_020 | TRUE |
| 0 | 0 | CHD3_dn_021 | TRUE |
| 1 | 0 | CHD3_dn_022 | FALSE |
| 0 | 0 | CHD3_dn_023 | TRUE |
| 0 | 0 | CHD3_dn_024 | TRUE |
| 0 | 0 | CHD3_dn_025 | TRUE |
| 0 | 0 | CHD3_dn_026 | TRUE |
| 0 | 0 | CHD3_dn_027 | TRUE |
| 1 | 0 | CHD3_dn_028 | FALSE |
| 1 | 0 | CHD3_dn_029 | FALSE |
| 0 | 0 | CHD3_dn_030 | TRUE |
| 1 | 0 | CHD3_dn_031 | FALSE |
| 0 | 0 | CHD3_dn_032 | TRUE |
| 1 | 0 | CHD3_dn_033 | FALSE |
| 0 | 0 | CHD3_dn_034 | TRUE |
| 0 | 0 | CHD3_dn_035 | TRUE |
| 0 | 1 | CHD3_inh_001 | FALSE |
| 1 | 1 | CHD3_inh_002 | TRUE |
| 0 | 1 | CHD3_inh_003 | FALSE |
| 0 | 1 | CHD3_inh_004 | FALSE |
| 0 | 1 | CHD3_inh_005 | FALSE |
| 0 | 1 | CHD3_inh_007 | FALSE |
| 0 | 1 | CHD3_inh_008 | FALSE |
| 0 | 1 | CHD3_inh_009 | FALSE |
| 0 | 1 | CHD3_inh_010 | FALSE |
| 0 | 1 | CHD3_inh_011 | FALSE |
| 0 | 1 | CHD3_inh_012 | FALSE |
| 1 | 1 | CHD3_inh_013 | TRUE |
| 1 | 1 | CHD3_inh_014 | TRUE |
| 1 | 1 | CHD3_inh_015 | TRUE |
| 1 | 1 | CHD3_inh_016 | TRUE |
| 1 | 1 | CHD3_inh_017 | TRUE |
| 0 | 1 | CHD3_inh_018 | FALSE |
| 0 | 1 | CHD3_inh_019 | FALSE |
| 0 | 1 | CHD3_inh_020 | FALSE |

| Cluster | Labels_reversed | Index | CORRECT_CLASSIFICATIE |
|---|---|---|---|
| 0 | 0 | CHD3_inh_001 | TRUE |
| 1 | 1 | CHD3_inh_001_p | TRUE |
| 0 | 0 | CHD3_inh_002 | TRUE |
| 1 | 1 | CHD3_inh_002_p | TRUE |
| 0 | 0 | CHD3_inh_003 | TRUE |
| 1 | 1 | CHD3_inh_003_p | TRUE |
| 1 | 0 | CHD3_inh_004 | FALSE |
| 1 | 1 | CHD3_inh_004_p | TRUE |
| 0 | 0 | CHD3_inh_005 | TRUE |
| 1 | 1 | CHD3_inh_005_p | TRUE |
| 0 | 0 | CHD3_inh_007 | TRUE |
| 1 | 1 | CHD3_inh_007_p | TRUE |
| 1 | 0 | CHD3_inh_008 | FALSE |
| 1 | 1 | CHD3_inh_008_p | TRUE |
| 1 | 0 | CHD3_inh_009 | FALSE |
| 1 | 1 | CHD3_inh_009_p | TRUE |
| 0 | 0 | CHD3_inh_010 | TRUE |
| 1 | 1 | CHD3_inh_010_p | TRUE |
| 0 | 0 | CHD3_inh_011 | TRUE |
| 1 | 1 | CHD3_inh_011_p | TRUE |
| 1 | 0 | CHD3_inh_012 | FALSE |
| 1 | 1 | CHD3_inh_012_p | TRUE |
| 0 | 0 | CHD3_inh_013 | TRUE |
| 0 | 1 | CHD3_inh_013_p | FALSE |
| 0 | 0 | CHD3_inh_014 | TRUE |
| 1 | 1 | CHD3_inh_014_p | TRUE |
| 0 | 0 | CHD3_inh_015 | TRUE |
| 1 | 1 | CHD3_inh_015_p | TRUE |
| 0 | 0 | CHD3_inh_016 | TRUE |
| 1 | 1 | CHD3_inh_016_p | TRUE |
| 0 | 0 | CHD3_inh_017 | TRUE |
| 1 | 1 | CHD3_inh_017_p | TRUE |
| 1 | 0 | CHD3_inh_018 | FALSE |
| 1 | 1 | CHD3_inh_018_p | TRUE |
| 0 | 0 | CHD3_inh_019 | TRUE |
| 1 | 1 | CHD3_inh_019_p | TRUE |
| 0 | 0 | CHD3_inh_020 | TRUE |
| 1 | 1 | CHD3_inh_020_p | TRUE |

**Figure S4 Grouped HPO features based on semantic similarity and clustering results per individual. A)** The semantic similarity between all the HPO terms used in this cohort was calculated using the Wang algorithm in the HPOsim package in R. HPO terms with at least a 0.5 similarity score were grouped and a new feature was created as a replacement, which was the sum of the grouped features. **B**) Individual HPO-based phenotypic clustering results for analyses comparing individuals with *de novo* (dn under ‘Index’) and inherited variants (inh under ‘Index’; left), and probands and carrier parents (indicated with a ‘p’ under ‘Index’; right). Index numbers match the family numbering of the cohort.

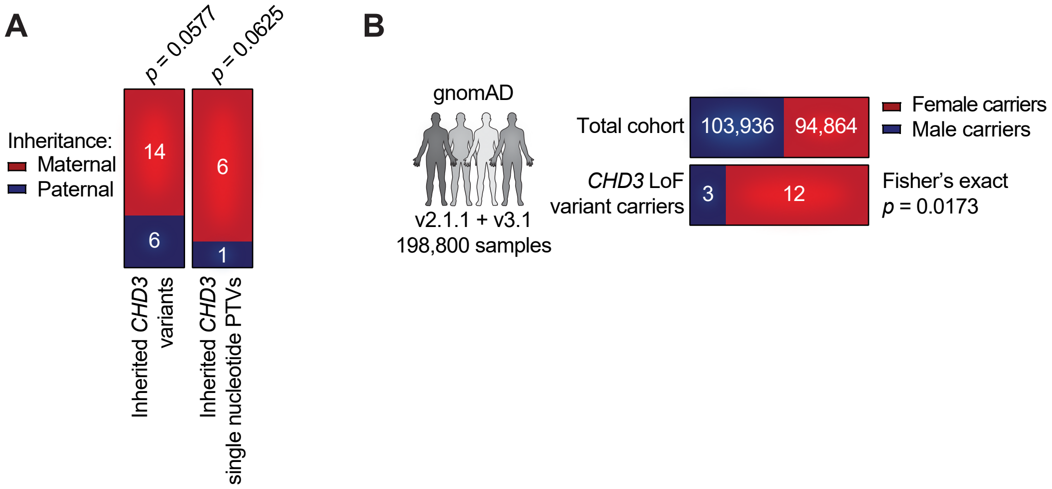


**Figure S5. Parental inheritance of inherited *CHD3* variants and sex distribution of *CHD3* loss-of-function variant carriers in healthy individuals. A**) Inheritance of all inherited *CHD3* variants (left), and of only inherited *CHD3* single nucleotide PTVs (right), in families with a proband with NDD (one-sided bionomial test with expected ratios of 0.5 for paternal inheritance and 0.5 for maternal inheritance. The *p*-value is the probability of the found number of maternally inherited cases or more). **B**) Sex distribution of the GnomAD cohort (top) and of *CHD3* LoF variant carriers in the GnomAD cohort (bottom; *p*-value based on a two-sided Fisher's exact test). Only carriers with stop-gain and frameshift variants were included. Carriers of first and last exon variants were excluded, as well as carriers of pLoF-flagged variants and mosaic cases.

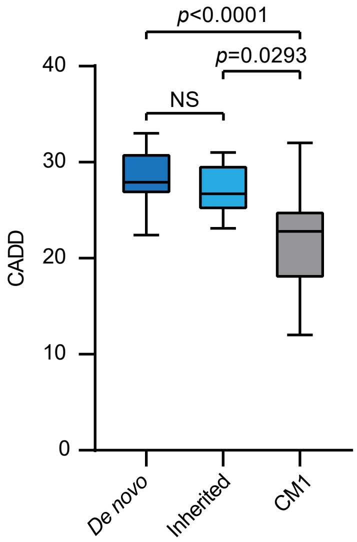


| *De novo* CHD3 missense variants | CADD v1.6 |
|---|---|
| p.Gln569Arg | 33 |
| p.His886Arg | 25.9 |
| p.Leu915Phe | 23.7 |
| p.Asn917Tyr | 28.6 |
| p.Glu921Lys | 29.8 |
| p.Phe944Tyr | 26.6 |
| p.Ser948Pro | 27.3 |
| p.Gly961Glu | 27.9 |
| p.Arg966Trp | 30 |
| p.Arg966Pro | 31 |
| p.Lys969Glu | 27.3 |
| p.Arg985Trp | 29.8 |
| p.Arg985Gln | 32 |
| p.Arg1121Pro | 31 |
| p.Thr1136Ile | 26.6 |
| p.Trp1158Arg | 27.9 |
| p.Asn1159Lys | 22.8 |
| p.His1161Arg | 24.9 |
| p.Arg1169Trp | 26.7 |
| p.His1171Arg | 26.6 |
| p.Arg1172Gln | 32 |
| p.Leu1080His | 28.5 |
| p.Arg1187Pro | 27.2 |
| p.Leu1236Pro | 28.5 |
| p.Arg1262Trp | 26.8 |
| p.Arg1342Gln | 31 |
| p.Arg1415Cys | 32 |
| p.Arg1881Leu | 31 |
| p.Ala1955Ser | 22.4 |
| **Mean** | 28.23 |
| **Standard deviation** | 2.81 |

| Inherited CHD3 missense variants | CADD v1.6 |
|---|---|
| p.Ser477Phe | 25.8 |
| p.Trp630Arg | 29.5 |
| p.Phe833Leu | 24.9 |
| p.Gly873Ser | 23.1 |
| p.Ile983Val | 25.1 |
| p.Pro1046Leu | 31 |
| p.Arg1342Gln | 31 |
| p.Arg1706Gln | 25.3 |
| p.Arg1759Gln | 29.9 |
| p.Glu1837Lys | 29.5 |
| p.Arg1888Gln | 27.6 |
| p.Lys1972Thr | 23.1 |
| **Mean** | 27.15 |
| **Standard deviation** | 2.95 |

| Rare CHD3 missense variants associated with CM1 | CADD v1.6 |
|---|---|
| p.Cys17Tyr* | 18.81 |
| p.Arg24Trp* | 22.8 |
| p.Ala25Val* | 16.31 |
| p.Lys114Thr | 19.58 |
| p.Arg271Gln | 25.7 |
| p.Arg337Trp | 25 |
| p.Tyr595Cys | 23.6 |
| p.Gly733Arg | 32 |
| p.Arg876Cys | 23.5 |
| p.Asp1358Glu | 16.97 |
| p.Arg1600Gln | 22.8 |
| p.Val1624Leu | 12.02 |
| p.Ala1937Thr | 17.83 |
| p.Lys1972Glu | 23.9 |
| p.Cys1997Tyr | 26.8 |
| **Mean** | 21.84 |
| **Standard deviation** | 4.97 |

**Figure S6. In silico prediction of pathogenicity scores for *CHD3* variants in NDD and CM1.** CADD scores (v1.6) for *de novo CHD3* variants in NDD, described in Snijders Blok et al. 2018 and Drivas et al. 2020, for inherited *CHD3* variants identified in the current study, and for rare missense variants associated with Chiari I malformation (CM1) described in Saddler et al. 2021 were obtained, and plotted as box plots, showing the median CADD scores for each group. CADD scores for *de novo* and inherited *CHD3* missense variants in NDD are significantly higher compared to CADD scores of missense variants associated with CM1 without NDD, while *de novo* and inherited *CHD3* missense variants in NDD do not have significantly different CADD scores (Kruskal-Wallis test followed by a Dunn's multiple comparisons test, NS: not significant). In the table, positions are with reference to sequence ENST00000380358.4 (NM_001005273.2), variants marked with an asterisk are with reference to sequence ENST00000481999.1.

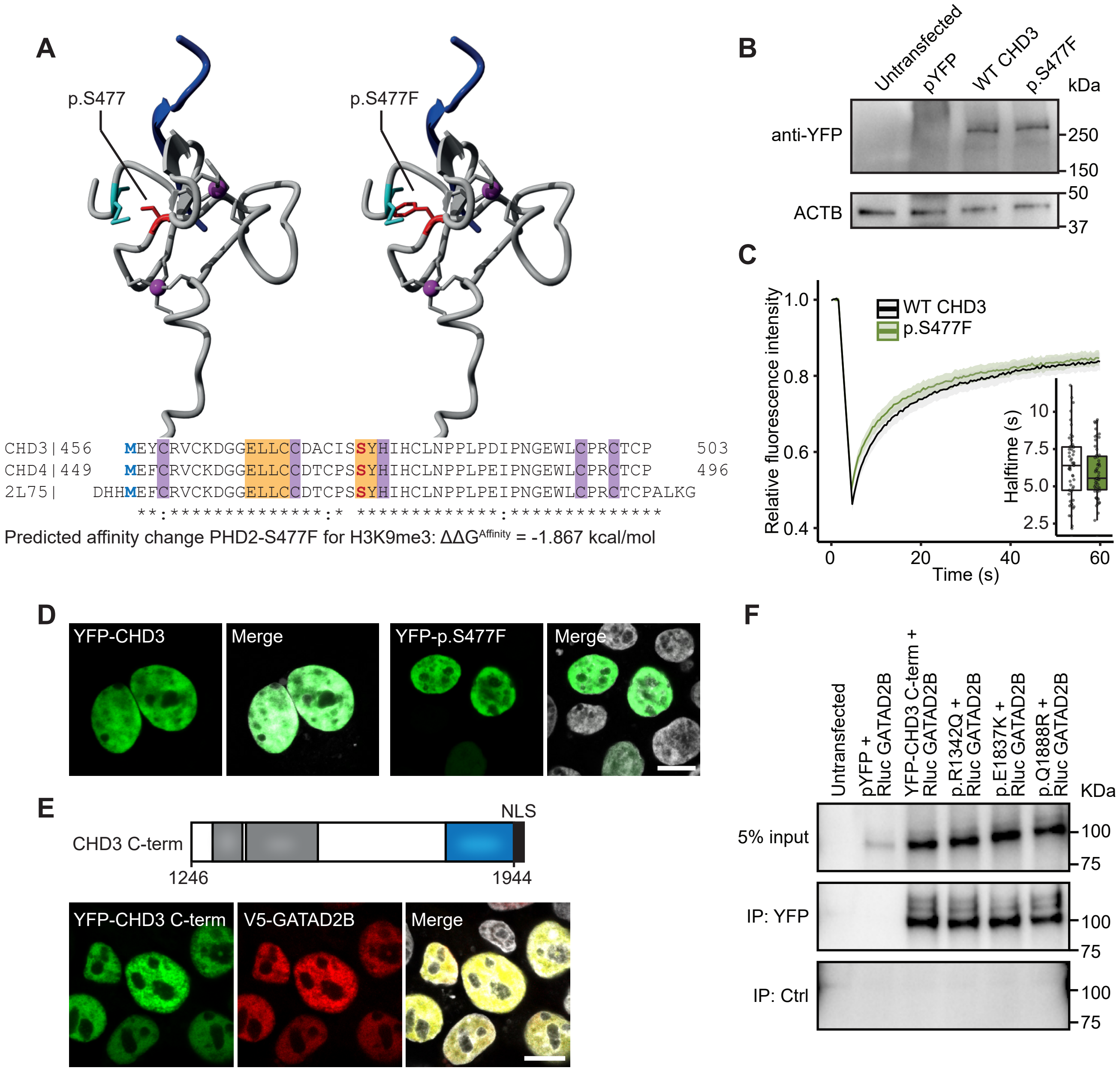


**Figure S7. Functional characterization of inherited *CHD3* missense variants. A**) Three-dimensional modeling of the CHD3 p.S477F variant in the PHD2 domain of CHD4 (PDB 2L75, 89% sequence identity with CHD3). In both the protein model and the alignment, the affected residue is depicted in red, and p.M456, that forms a hydrogen bond with p.S477, is shown in cyan. Zinc metals, and zinc binding residues are purple, and H3K9me3 is shown in dark blue. Residues that form a beta-sheet are shaded in yellow in the alignment. The effect of p.S477F on binding of PHD2 to H3K9me3 was assessed using the mCSM-PPI2 machine learning tool, with $\Delta\Delta G^{Affinity} = \Delta\Delta G_{wt-mt} = \Delta G_{wild-type} - \Delta G_{mutant}$. The p.S477F variant is predicted to impact CHD3 histone binding. **B**) Immunoblot of whole-cell lysates expressing YFP-tagged CHD3 and the p.S477F variant probed with anti-EGFP antibody, showing bands at the expected molecular weight of ~260 kDa. The blot was probed for ACTB to ensure equal protein loading. **C**) FRAP experiments to assess the dynamics of CHD3 chromatin binding in live cells. Graph shows the mean recovery curves ± 95% C.I. recorded in HEK293T/17 cells expressing YFP-CHD3 fusion proteins. Right corner, box plot of the halftime based on single-term exponential curve fitting of individual recordings ($n$ = 55 nuclei from three independent experiments, no significant difference, Student's t-test). The protein mobility of the p.S477F variant in the nucleus is not significantly different from WT protein, suggesting no loss of chromatin binding. **D**) Direct fluorescence micrographs of HEK293T/17 cells expressing YFP-CHD3 fusion proteins (green). Nuclei were stained with Hoechst 33342 (white). Scale bar = 10 µm. The p.S477F variant localized to the nucleus similar to WT protein. **E**) Top: Schematic of the cCHD3-NLS construct used for protein-interaction assays, encoding residues 1246-1944 including the CHDCT2 domain (blue), and an SV40-NLS appended at the C-terminus (black). Bottom: Direct fluorescence micrographs of HEK293T/17 cells co-expressing YFP-CHD3 (green) and V5-GATAD2B (red). Nuclei were stained with Hoechst 33342 (white). Scale bar = 10 µm. **F**) Co-immunoprecipitation assay to study the CHD3-GATAD2B interaction. YFP-tagged C-terminal truncations of CHD3 (WT, p.R1342Q, p.E1837K, p.R1888Q) were coexpressed with Rluc-GATAD2B in HEK293T/17 cells. YFP-fusion proteins were immobilized on αGFP affinity beads and used as baits to pull down the coexpressed Rluc-GATAD2B. Lysates of untransfected cells and cells co-transfected with pYFP and Rluc-GATAD2B were used as negative controls. As binding control, lysates were incubated with deactivated beads. All tested CHD3 C-terminal variants were still able to pull-down GATAD2B similar to WT protein, suggesting that they do not disrupt the CHD3-GATAD2B interaction.

**Table S5. Primers to amplify the region that carries the *CHD3* p.W1158* stop-gain variant to test for NMD.**

| | |
|---|---|
| CHD3-NMD-W1158*-F | TCGGTTTAATGCTCCTGGGG |
| CHD3-NMD-W1158*-R | ACAACCAGGTGTGTCAGCAT |

**Table S6. Primers to clone the CHD3 C-terminal construct and GATAD2B.** Underscored italic sequence marks the restriction sites (*HindIII*/*BamHI*) and the bold sequence is an SV40 NLS sequence that was added to ensure nuclear localization of the encoded protein.

| | |
|---|---|
| CHD3-Cterm-cloning-F1 | GAGGGG<u>*AAGCTT*</u>AAAGGAGGAGGACAGCAGTGT |
| CHD3-Cterm-cloning-R1+NLS | TGCAGG<u>*GGATCC*</u>TCA**GACCTTGCGCTTCTTCTT**CGGGGCCAGGGCCCCTACGGGTG |
| GATAD2B-cloning-F1 | <u>*GGATCC*</u>TGGATAGAATGACAGAAGATGC |
| GATAD2B-cloning-R1 | <u>*TCTAGA*</u>TTATTTCTGTCCACTGATGG |

**Table S7. Primers used for site-directed mutagenesis.**

| | |
|---|---|
| CHD3 p.S477F F | GACAATGAATGTGGTAGAAGGAGATGCACGCGTCA |
| CHD3 p.S477F R | TGACGCGTGCATCTCCTTCTACCACATTCATTGTC |
| CHD3 p.R1342Q F | CTTGCTTGCGAACCTGCTTGCCCTTGCCT |
| CHD3 p.R1342Q R | AGGCAAGGGCAAGCAGGTTCGCAAGCAAG |
| CHD3 p.E1837K F | GGCCAGGCACTTGGCCTCGGCGA |
| CHD3 p.E1837K R | TCGCCGAGGCCAAGTGCCTGGCC |
| CHD3 p.R1888Q F | GATGGGGGGTATTTGGGACAGCGTGGC |
| CHD3 p.R1888Q R | GCCACGCTGTCCCAAATACCCCCCATC |
