## Supplemental notes for "Inherited variants in *CHD3* demonstrate variable expressivity in Snijders Blok-Campeau syndrome": 20210924-SupplementalNotes2.pdf

### Supplemental Notes 2

#### 3D-protein modeling of inherited CHD3 ATPase-Helicase variants

We modeled the protein structure of the ATPase-Helicase domain of CHD3 in interaction with the DNA using the homology modeling script in the WHAT IF [1] & YASARA [2] Twinset with standard parameters. As a template, we used PDB file 6RYR which contains the human Nucleosome-CHD4 complex structure of a single copy of CHD4 [3]. We performed mutation analysis on previously published *de novo* CHD3 ATPase-Helicase domain missense variants [4; 5], and on four inherited CHD3 ATPase-Helicase domain missense variants (p.F833L, p.G873S, p.I983V, p.P1046L; Supp. N2 - Figure 1).

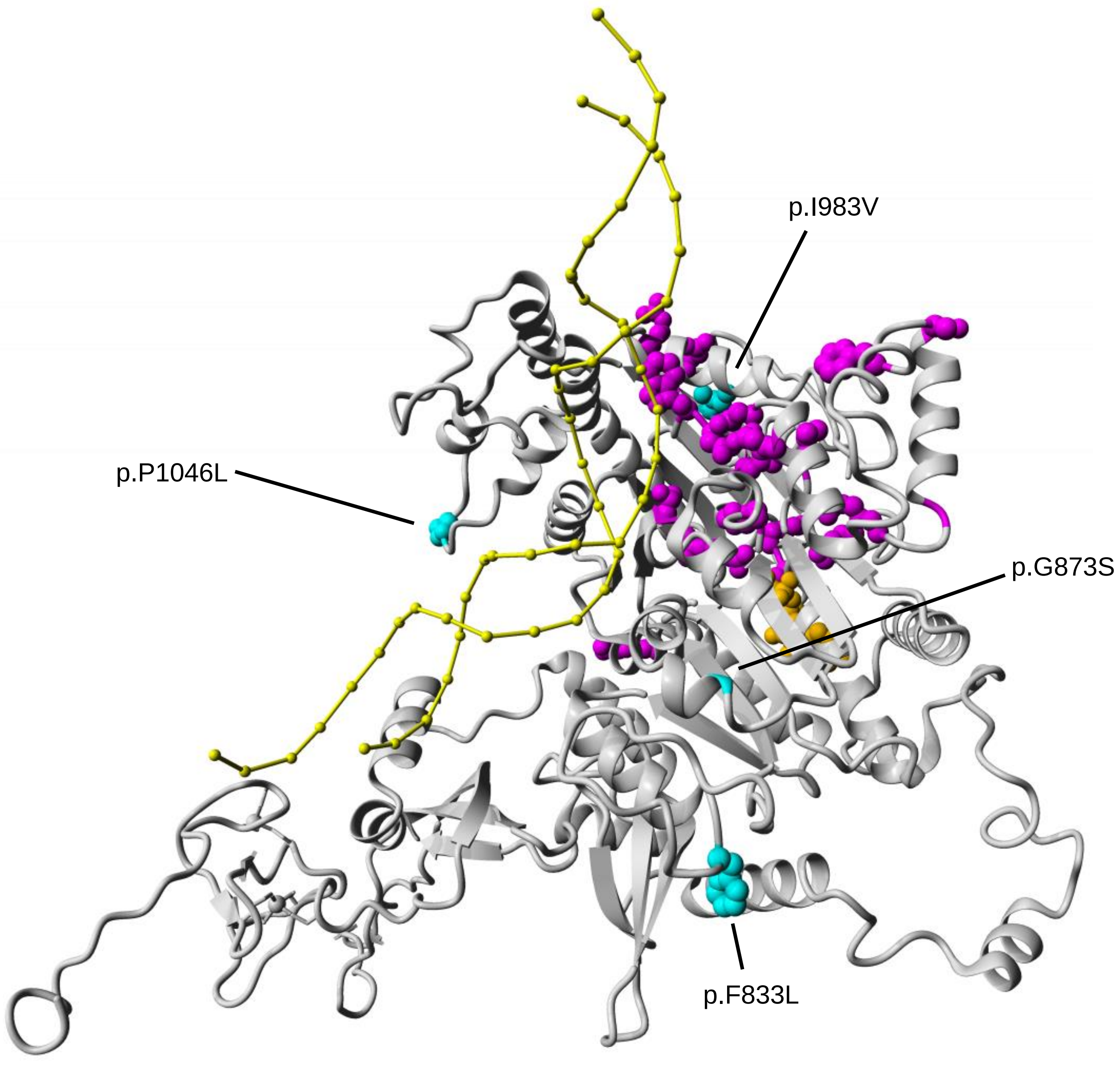


**Supp. N2 - Figure 1. Overview of *de novo* and inherited CHD3 variants in the ATPase-Helicase domain.**

3D model of the CHD3 ATPase-Helicase domain (homology model based on PDB: 6RYR; gray) in interaction with the DNA (yellow). ATP is depicted in orange. *De novo* CHD3 variants in the ATPase Helicase domain are shown in magenta, and cluster at DNA- and ATP-binding sites. The inherited CHD3 variants in this domain are depicted in cyan, and seem further away from DNA- and ATP-binding sites.

**Mutation analysis**

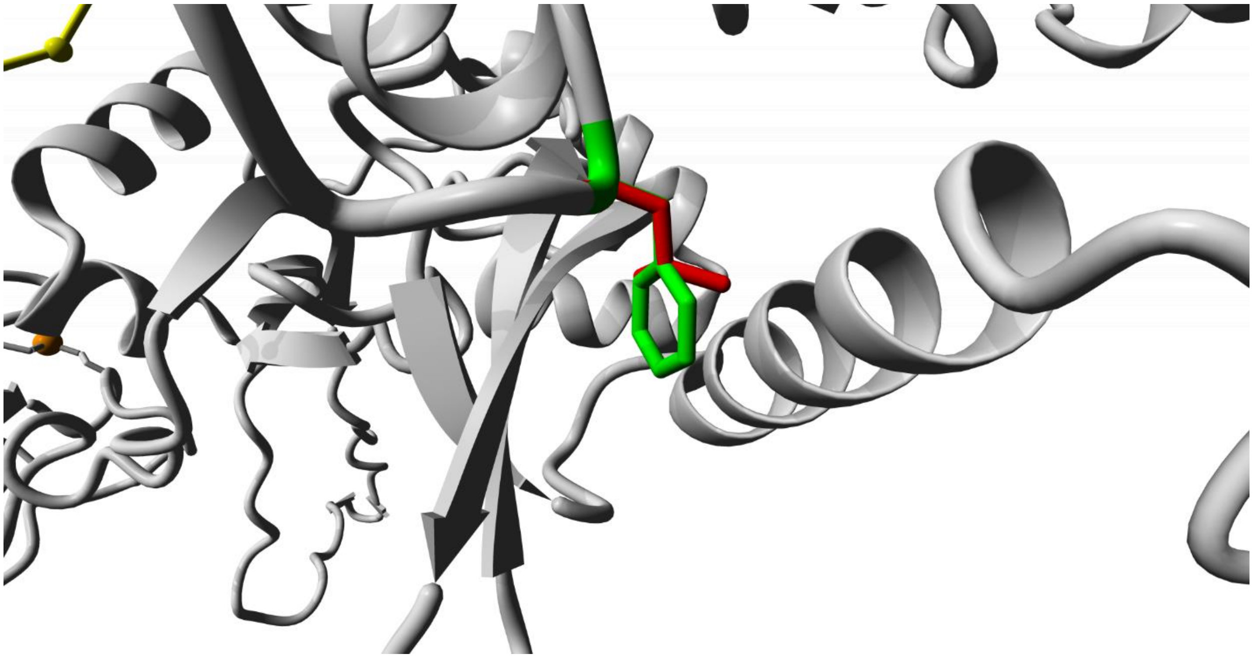

**Supp. N2 - Figure 2.** Close-up of the p.F833L variant. The CHD3 protein is colored in grey and the DNA is shown in yellow. The wild-type (F) and mutant (L) sidechains are shown in green and red respectively.

**p.F833L (Supp. N2 - Figure 2)**: This change concerns a semiburied residue. Leucine has a smaller sidechain and therefore some hydrophobic interactions will be lost leading to a slight destabilization of this area. This could affect the interaction with either the DNA or another protein, but only slightly.

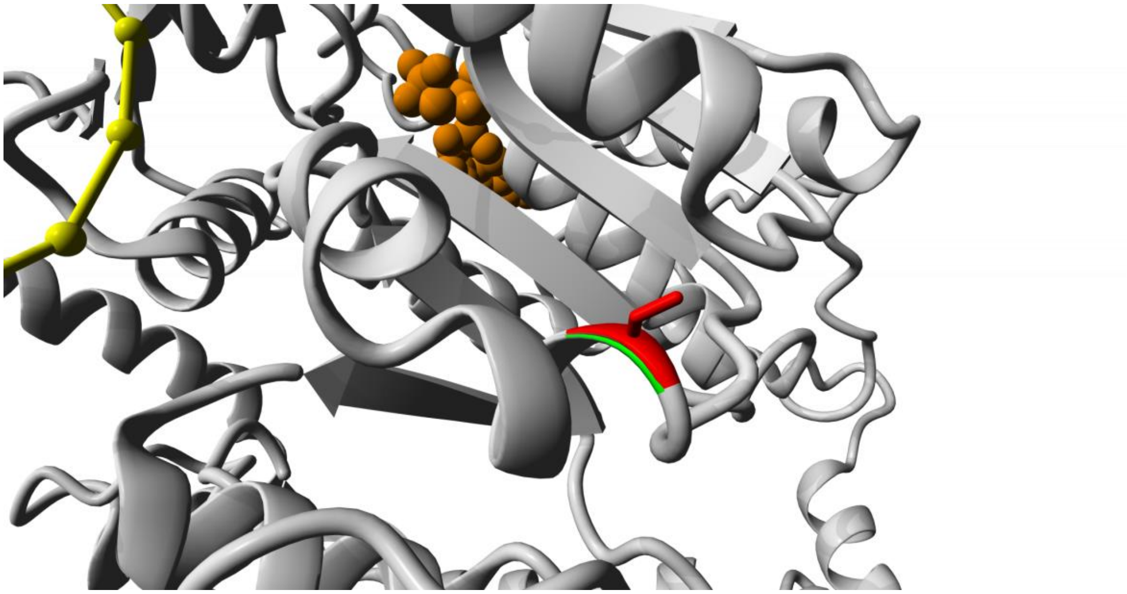

**Supp. N2 - Figure 3.** Close-up of the p.G873S variant. The CHD3 protein is colored in grey, the ATP in orange and the DNA is shown in yellow. The wild-type (G) and mutant (S) sidechains are shown in green and red respectively.

**p.G873S (Supp. N2 - Figure 3)**: Introduces a slightly bigger sidechain that is also known to interact with DNA. Since the residue is semiburied, only a small destabilization would be expected.

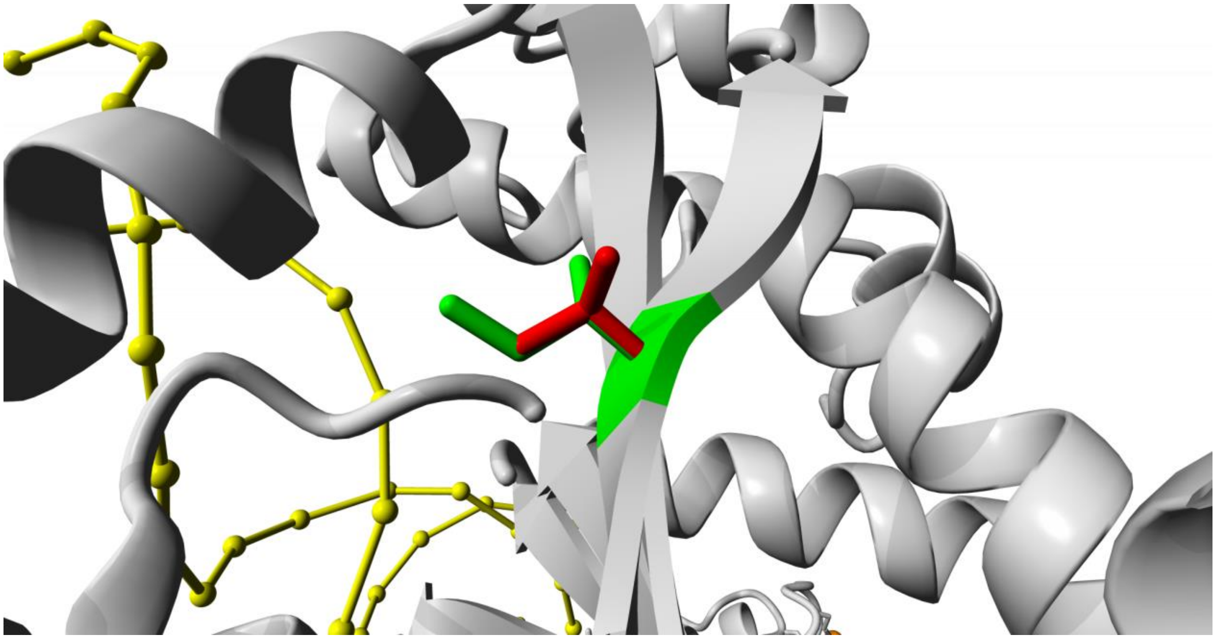

**Supp. N2 - Figure 4.** Close-up of the p.I983V variant. The CHD3 protein is colored in grey and the DNA is shown in yellow. The wild-type (I) and mutant (V) sidechains are shown in green and red respectively.

**p.I983V (Supp. N2 - Figure 4)**: Also a semiburied residue and located far away from the DNA. The loss of some hydrophobic interactions made by the sidechain does not seem to have many consequences.

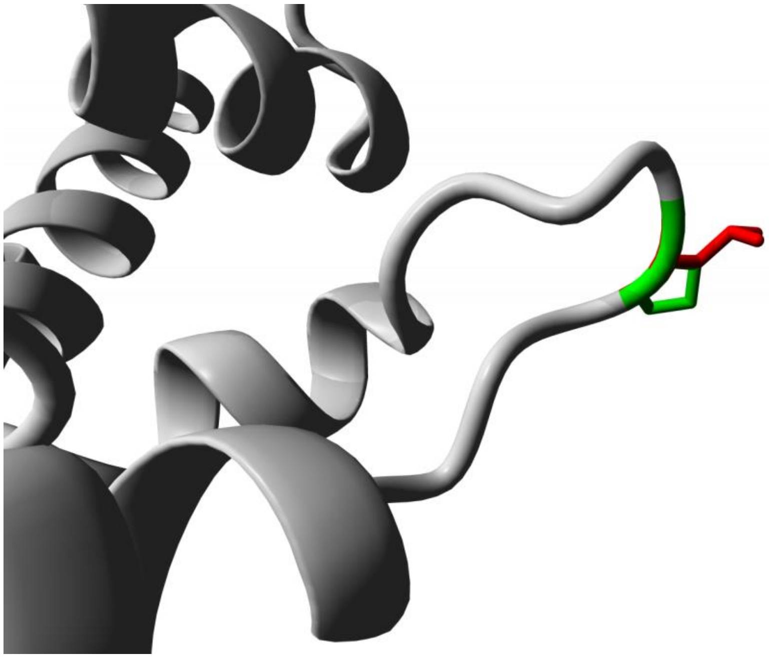

**Supp. N2 - Figure 5.** Close-up of the p.P1046L variant. The CHD3 protein is colored in grey and the DNA is shown in yellow. The wild-type (P) and mutant (L) sidechains are shown in green and red respectively.

**p.P1046L (Supp. N2 - Figure 5)**: Located in a clear and possibly flexible surface loop, where the new and larger leucine sidechain easily fits. The stable structure of the loop caused by proline will be lost. The function of this loop is unclear, but might be necessary for interaction with other proteins.
