## Supplemental notes for "Inherited variants in *CHD3* demonstrate variable expressivity in Snijders Blok-Campeau syndrome": 20210924-SupplementalNotes3.pdf

**Supplemental Notes 3**

**Population-level genetic analyses in UK Biobank**

### 1. Materials and methods

#### 1.1 Data

##### *1.1.1 UK Biobank*

The research described in this section has been conducted using the UK Biobank (UKB) Resource under application 16066, with Clyde Francks as the principal applicant. This is a general adult population cohort [1]. The data collection in the UKB, including the consent procedure, has been described elsewhere [2]. Informed consent was obtained by the UKB for all participants. We made use of imaging-derived phenotype data generated by an image-processing pipeline developed and run on behalf of the UKB [3; 4].

##### *1.1.2 Whole exome sequencing data*

Whole-exome sequencing (WES) of approximately 200,000 individuals was performed through the UKB Exome Sequencing Consortium according to protocols described elsewhere [5; 6] (https://www.ukbiobank.ac.uk/media/cfulxh52/uk-biobank-exome-release-faq_v9-december-2020.pdf). Briefly, exomes were captured with the IDT xGen Exome Research Panel v1.0 including supplementary probes, targeting 39 Mbp of the human genome. Sequencing was performed on the Illumina NovaSeq 6000 platform and data was processed using the OQFE protocol [5]: raw reads were mapped to a GRCh38 reference, retaining all supplementary alignments, and duplicate reads were marked. Variants were called, restricted to exome capture regions plus the 100 basepairs flanking each capture target, resulting in a genomic variant-call file (gVCF) per sample. gVCFs were then merged into multi-sample project-level VCF (pVCF) files. We downloaded the multi-sample pVCF files and removed variants outside sequencing target regions defined by the UKB, as sequencing quality standards for variants outside these regions were not assessed. We then extracted variants in the *CHD3* gene region, which we defined as chr17:7884796-7912760 (genome build

GRCh38/hg38) or chr17:7788124-7816078 (GRCh37/hg19), yielding 1,056 variant sites. Variants in the *CHD3* region were annotated using snpEff v5.0 (build 2020-10-04) [7]. In addition to default snpEff annotations, we annotated variants with SIFT4G [8], PolyPhen2-HDIV [9] and CADD [10] scores derived from dbNSFP (version 4.1a) [11]. SIFT4G scores ranged from 0-1, with smaller scores indicating a more likely damaging effect. Within dbNSFP, variants with a SIFT4G score < 0.05 were labelled as 'damaging'. PolyPhen2-HDIV scores ranged from 0 to 1, with higher scores indicating a more likely damaging effect. Within dbNSFP, variants with a PolyPhen-2 score [0.957-1] were labeled as 'probably damaging'. CADD scores were presented as Phred-like ($log_{10}$-derived) rank scores based on the distribution of all CADD scores across the genome, with higher Phred-scores indicating higher predicted deleteriousness.

#### 1.2 Association of rare variants with educational qualifications, fluid intelligence score and intracranial volume

##### *1.2.1 Phenotype data*

We selected three main phenotypes for association testing with rare missense variants in *CHD3*: 'Fluid intelligence score' (data field ID 20016), 'Qualifications' (data field ID 6138) and 'Volume of EstimatedTotalIntraCranial (whole brain)' (data field ID 26521). For each main phenotype, additional phenotypes were selected to use as covariates in the association tests (Supp. N3 - Table 1). Fluid intelligence was recorded at multiple instances, and for each individual the data from the first available instance was used. Qualifications was also reported at multiple instances and had six possible answering options (1: College or University degree, 2: A levels/AS levels or equivalent, 3: O levels/GCSEs or equivalent, 4: CSEs or equivalent, 5: NVQ or HND or HNC or equivalent, and 6: Other professional qualifications (e.g. nursing, teaching, etc.)), with multiple possible answers per instance. To make this phenotype suited for association testing, categories were merged to create a binary phenotype: one group comprised individuals for which the highest reported qualification was option 1 or 2 ('high' educational qualification), and the other group comprised individuals for which the highest reported qualification was option 3, 4 or 5 ('low' educational qualification). Because option 6 (other professional qualifications) can refer to diverse educational levels, we excluded individuals who only reported this option as their highest qualification. Furthermore, we excluded individuals if they were inconsistently assigned to both ‘high’ and ‘low’ categories across instances. In all datasets, we included the first 10 principal components (PCs) based on common variant data that reflect genetic ancestry [1], and the exome sequencing batch (a binary variable indicating whether an individual was sequenced in the first batch of ~50,000 individuals or in the second batch of ~150,000 individuals) as additional covariates. We made a separate dataset for each of the three main phenotypes, including only individuals with available WES data.

##### *1.2.2 Sample-level filtering*

In each of the three datasets, individuals with missing data for one or more covariates were excluded. We additionally excluded individuals with discordant phenotypic and genetically inferred sex (as reported in the exome sequencing data plink .fam file), as well as those not in the white-British ancestry subset as defined by the UKB based on self-reported ancestry and clustering in the first six genetic PCs [1]. In pairs of individuals defined as third-degree relatives or closer (kinship coefficient > 0.0442) by the UKB [1], we randomly removed one individual of a pair, prioritizing removal of individuals related to multiple others.

##### *1.2.3 WES data filtering*

Following sample-level filtering, we applied genotype- and variant site-level hard-filtering in each phenotype dataset using vcftools 0.1.17 [12] and previously published thresholds [6; 13]. Genotype-level filtering changed genotypes with a low approximate read depth (DP < 7 for single-nucleotide variant [SNV] sites and DP < 10 for insertion/deletion [INDEL] sites) and/or low genotype quality (GQ < 20) to no-call. Variant-level filtering removed sites labeled as ‘monoallelic’, sites with low average genotype quality across individuals (average GQ < 35), a high missingness rate (> 0.12), a minor allele count (MAC) of zero, and/or a low allele balance (AB < 0.15 for SNV sites and < 0.20 for INDEL sites, calculated using GATK v4.1.9.0 [14]).

##### *1.2.4 Region-based association test of missense variants*

For region-based association testing, we removed multi-allelic variants and converted data to plink binary format using plink v1.90b6.9 [15]. We then used MetaDome to select missense variants at mutation-intolerant locations in functional domains of *CHD3* [16]. Only variants in the three best characterized transcripts of *CHD3* (Ensembl transcript IDs: ENST00000330494, ENST00000358181, ENST00000380358), a predicted ‘damaging’ (SIFT4G) or ‘probably damaging’ (PolyPhen2-HDIV) effect and a CADD PHRED score > 25, and a minor allele frequency (MAF) ≤ 1% were included. Optimized sequence kernel association testing (SKAT-O), implemented in the R package *SKAT* [17], was used to capture the combined association of the selected rare missense variants in *CHD3* with the three selected phenotypes. The SKAT-O test was designed to maximize power by optimally combining a burden and sequence-kernel association test (SKAT), making it both suitable under scenarios where rare variants within *CHD3* are associated with corresponding direction of effect, as well as in the presence of null effects or opposing effect directions. Variants were weighted for MAF using the default beta(1,25) density function implemented in SKAT, and null models for each phenotype included the covariates as shown in Supp. N3 -Table 1. For the educational qualification phenotype, we used an improved ‘robust’ version of the SKAT-O method for association testing of binary phenotypes [18].

##### *1.2.5 Phenotypes of putative loss-of-function variant carriers*

We extracted protein coding variants with a high putative impact in the three best characterized transcripts (Ensembl transcript IDs: ENST00000330494, ENST00000358181, ENST00000380358) of *CHD3* from the remaining variants after filtering. Individuals carrying one or more putative loss-of-function (LoF) variants were identified in each of the three phenotype datasets. We then applied linear regression analysis to assess differences in fluid intelligence score and intracranial volume between LoF-carriers and controls (non-carriers), and projected LoF-carrier phenotypes on the phenotype distribution of controls. For educational qualification, we used logistic regression to assess group differences. All regression analyses included the same set of covariates for each main phenotype as in the association test of missense variants (listed in Supp. N3 – Table 1).

##### 1.3 Common variant association of *CHD3* with head circumference and intracranial volume

We used association results of SNPs located inside the *CHD3* gene region, derived from previously published genome-wide association meta-analysis summary statistics of head circumference (N ≤ 18,881), and head circumference combined with intracranial volume (N ≤ 45,458), in child- and adulthood [19]. This study included both common and low-frequency genetic variants. In addition, we obtained results from a smaller genome-wide association meta-analysis on infant head circumference (N ≤ 10,768) [20]. For each study, we calculated gene p-values reflecting the common variant association of *CHD3* with HC and ICV using the default ‘snp-wise=mean’ analysis model in MAGMA (v1.09a) [21]. We applied Bonferroni correction for three included genome-wide studies when assessing significance.

#### 2. Results

##### 2.1 Region-based association testing of missense variants

For the three main phenotypes, we selected individuals with available WES data and a valid reported phenotype. We then performed sample-level quality control, leaving 120,596 individuals for association analysis of educational qualification, 77,998 for fluid intelligence and 18,254 for intracranial volume (Supp. N3 - Table 2). We extracted genotype data for the individuals in each dataset and performed genotype- and variant-level filtering on 1,056 variant sites present in the defined *CHD3* region (Supp. N3 - Table 3). From variants remaining after filtering, we extracted missense variants on mutation intolerant locations in functional domains of *CHD3*. 47 selected variants overlapped with the post-QC variants in the three datasets, with 43 variants present in the educational qualification dataset, 37 variants in the fluid intelligence dataset, and 13 variants in the intracranial volume dataset. SKAT-O analysis revealed no significant association of missense variants in *CHD3* with educational qualification (p = 0.35), fluid intelligence score (p = 0.36) and intracranial volume (p = 0.57) (Supp. N3 - Table 4).

##### 2.2 Phenotypes of putative LoF variant carriers

We only identified heterozygous carriers of putative LoF variants. There was no significant effect of LoF-carrier group on educational qualification ($z$ = 0.427, p = 0.67) and fluid intelligence score ($t$ = 0.189, p = 0.85). Intracranial volumes of four putative LoF variant carriers were higher than the average in non-carriers, with a nominally significant group effect ($t$ = 2.37, p = 0.018) (Supp. N3 - Table 5, Supp. N3 - Figure 1).

##### 2.3 Common variant association of *CHD3* with head circumference and intracranial volume

Gene-based analysis in MAGMA revealed no significant associations of common variants in *CHD3* with head circumference, although the combined analysis of head circumference with intracranial volume suggested a subtle association (Supp. N3 - Table 6).

#### 3. Tables and figures

**Supp. N3 - Table 1: UK Biobank phenotypes selected for association testing**

| **Fluid intelligence score** | | | |
|---|---|---|---|
| **Field ID** | **Phenotype** | **Instance** | **Note** |
| 31 | Sex | 0 | |
| *20016* | *Fluid intelligence score* | 0, 1, 2 or 3 | For each individual, data from the first available instance was selected. |
| 21003 | Age when attended assessment centre | 0, 1, 2 or 3 | For each individual, age was selected from the same instance as the one from which fluid intelligence score was obtained. |
| 22009 | Genetic principal components 1-10 | 0 | |
| - | Exome batch | - | Binary variable indicating whether individuals were sequenced in the first (50k) or second (150k) batch. |

| **Educational qualifications** | | | |
|---|---|---|---|
| **Field ID** | **Phenotype** | **Instance** | **Note** |
| 31 | Sex | | |
| 34 | Year of birth | | |
| *6138* | *Qualifications* | 0, 1, 2 or 3 | |
| 22009 | Genetic principal components 1-10 | 0 | |
| - | Exome batch | - | Binary variable indicating whether individuals were sequenced in the first (50k) or second (150k) batch. |

| **Intracranial volume** | | | |
|---|---|---|---|
| **Field ID** | **Phenotype** | **Instance** | **Note** |
| 31 | Sex | 0 | |
| 54 | UK Biobank assessment centre | 2 | |
| 21003 | Age when attended assessment centre | 2 | |
| 22009 | Genetic principal components 1-10 | 0 | |
| 25734 | Inverted signal-to-noise ratio in T1 | 2 | |
| 25735 | Inverted contrast-to-noise ratio in T1 | 2 | |
| 25756 | Scanner lateral (X) brain position | 2 | |
| 25757 | Scanner transverse (Y) brain position | 2 | |
| 25758 | Scanner longitudinal (Z) brain position | 2 | |
| *26521* | *Volume of EstimatedTotalIntraCranial (whole brain)* | 2 | |
| - | Exome batch | - | Binary variable indicating whether individuals were sequenced in the first (50k) or second (150k) batch. |

| | | | |
|---|---|---|---|
| - | Nonlinear age | - | Calculated from 'age when attended assessment centre'. |

*The main phenotype for each analysis is highlighted in italic, other phenotypes were included as covariates. The Field ID column shows the phenotype identifier in the UK Biobank. Instance reflects the assessment centre visit(s) from which the recorded phenotype was used. 0: First assessment center visit | 1: Follow-up assessment center visit | 2: First imaging visit | 3: Second imaging visit. For some phenotypes, data from multiple instances was combined (see methods section).*

**Supp. N3 - Table 2: Overview sample-level filtering**

| Filtering step/Dataset | Qualifications | Fluid intelligence score | Intracranial volume |
|---|---|---|---|
| Available main phenotype before filtering | 154,065 | 100,300 | 21,727 |
| Missing data in one or more covariates | 133 | 95 | 383 |
| Discordant reported and genetic sex | 35 | 36 | 9 |
| Not in white British ancestry subset | 26,161 | 17,736 | 2,853 |
| Third-degree or higher relatedness to other individual(s) in dataset | 7,277 | 4,535 | 291 |
| Unique individuals flagged for removal | 33,469 | 22,302 | 3,473 |
| Individuals left after filtering | 120,596 | 77,998 | 18,254 |
| Female | 65,458 | 41,852 | 9,669 |
| Male | 55,138 | 36,146 | 8,585 |
| Age (mean) [range] | - | 58.25 [40-81] | 63.24 [45-81] |
| Year of birth (median) [range] | 1952 [1936-1970] | - | - |

**Supp. N3 - Table 3: Overview of WES data filtering**

| Filtering step/Dataset | Qualifications | Fluid intelligence score | Intracranial volume |
|---|---|---|---|
| Variant sites in target regions pre-filtering | 1,056 | 1,056 | 1,056 |
| Monoallelic sites | 15 | 15 | 15 |
| Low average genotype quality (average GQ < 35) | 2 | 2 | 2 |
| High missingness rate (> 0.12) | 6 | 6 | 7 |
| Minor allele count of zero (MAC = 0) | 324 | 457 | 785 |
| Low allele balance (SNV sites < 0.15, INDEL sites < 0.20) | 0 | 0 | 0 |
| Unique variant sites removed | 345 | 476 | 802 |
| Variant sites post-filtering | 711 | 580 | 254 |
| Ts-Tv ratio pre-filtering | 4.08 | 4.08 | 4.08 |
| Ts-Tv ratio post-filtering | 4.96 | 4.44 | 5.68 |
| Multiallelic sites removed prior to region-based testing | 88 | 77 | 43 |
| Missense variants at intolerant sites in functional regions of *CHD3* | 43 | 37 | 13 |

**Supp. N3 - Table 4: Region-based association test results**

| Phenotype | P-value | Q | Variants | MAC |
|---|---|---|---|---|
| Qualifications | 0.35 | 9977.66 | 43 | 197 |
| Fluid intelligence score | 0.36 | 53090.60 | 37 | 135 |
| Intracranial volume | 0.57 | 9789.92 | 13 | 33 |

*P-value: SKAT-O association test p-value | Q: SKAT-O test Q value (test statistic) | Variants: number of variants tested within CHD3 in association analysis | MAC: Total/Gene minor allele count across all tested variants.*

**Supp. N3 - Table 5: Main phenotypes for LoF variant carriers versus controls**

| Phenotype | N total | N LoF carriers | Frequency controls | Frequency LoF carriers |
|---|---|---|---|---|
| Qualifications | 120,596 | 25 | $N_{High}$ = 66,941<br>$N_{Low}$ = 53,630 | $N_{High}$ = 15<br>$N_{Low}$ = 10 |
| | | | **Mean (SD) controls** | **Mean (SD) LoF carriers** |
| Fluid intelligence score | 77,998 | 14 | 6.26 (2.09) | 6.42 (1.50) |
| Intracranial volume | 18,254 | 4 | 1549140 (150494) | 1740448 (82451) |

*N total: total sample size for phenotype | N LoF carriers: The number of LoF variant carriers in the phenotype dataset | For qualifications, the frequency of controls and LoF variant carriers in the 'high' and 'low' groups is shown. For fluid intelligence and intracranial volume, the phenotype means (SD) are shown for controls and LoF variant carriers.*

**Supp. N3 - Table 6: Gene-level common variant associations of *CHD3* with head circumference and intracranial volume**

| GWAS dataset | N SNPs | N | ZSTAT | P |
|---|---|---|---|---|
| Haworth et al. – Head circumference | 64 | 15564 | 0.826 | 0.20 |
| Haworth et al. – Head circumference + intracranial volume | 65 | 31166 | 1.68 | 0.047 |
| Taal et al. – Infant head circumference | 20 | 10727 | 0.693 | 0.24 |

*N SNPs: Number of SNPs included in analysis | N: (average) sample size | ZSTAT: Z-statistic of the gene in gene-level analysis | P: Gene p-value*

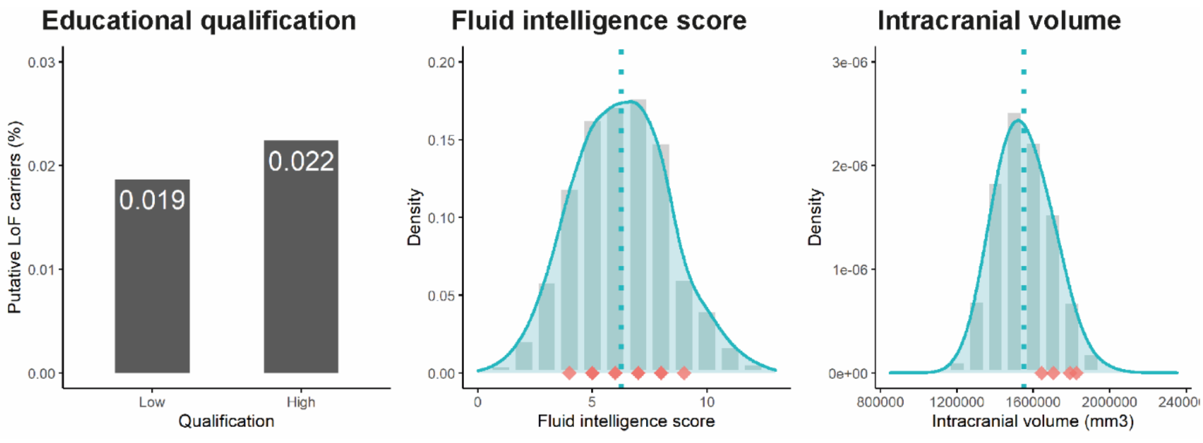


**Supp. N3 - Figure 1: Phenotype frequencies and distributions for putative loss-of-function variant carriers and non-carriers.** The bar plot for educational qualification shows the percentage of LoF variant carriers among all individuals in 'low' and 'high' educational qualifications groups. For fluid intelligence score and intracranial volume, phenotype distributions are shown for controls, and phenotype values for individual LoF carriers are projected on the distributions (red diamonds).
